# A multi-scale model to evaluate airport wastewater surveillance and ICU genomic monitoring for pandemic preparedness

**DOI:** 10.64898/2026.02.27.26347250

**Authors:** Benjamin K. Reddy, Joseph L.-H. Tsui, Kieran O. Drake, Guillaume St-Onge, Jessica T. Davis, Cathal Mills, Jake Dunning, Isaac I. Bogoch, Samuel V. Scarpino, Samir Bhatt, Oliver G. Pybus, Andrew Rambaut, Matthew J. Wade, Thomas Ward, Meera Chand, Erik M. Volz, Alessandro Vespignani, Moritz U.G. Kraemer

**Affiliations:** Pandemic Sciences Institute, University of Oxford, United Kingdom; Department of Statistics, University of Oxford, United Kingdom; Intelligent Earth Centre for Doctoral Training, University of Oxford, Oxford, United Kingdom; Department of Biology, University of Oxford, United Kingdom; Centre for Global Infectious Disease Analysis and Department of Infectious Disease Epidemiology, School of Public Health, Imperial College London, London, United Kingdom; Laboratory for the Modeling of Biological and Socio-technical Systems, Northeastern University, Boston, MA, USA; The Roux Institute, Northeastern University, Portland, ME, USA; Network Science Institute, Northeastern University, Boston, MA, USA; Department of Medicine, Division of Infectious Diseases, University of Toronto, Toronto, Canada; Department of Public Health and Health Sciences, Bouvé College of Health Sciences, Northeastern University, Boston, MA, USA; Institute for Experiential AI, Northeastern University, Boston, MA, USA; Santa Fe Institute, Santa Fe, NM, USA; Section of Epidemiology, Department of Public Health, University of Copenhagen, Copenhagen, Denmark; Pioneer Centre for Artificial Intelligence University of Copenhagen, Copenhagen, Denmark; Institute of Ecology and Evolution, University of Edinburgh, Edinburgh, UK; UK Health Security Agency, London, UK; Institute for Scientific Interchange Foundation, Turin, Italy

## Abstract

Increasing human mobility and population connectivity have intensified the risks of global pathogen spread, while concurrent shifts in human demographic patterns, ecological factors, and climatic conditions have altered the global landscape of this risk. Genomic surveillance can serve as a critical tool for early detection of emerging pathogen threats; however, challenges remain in deciding where to monitor, in understanding trade-offs among surveillance modalities, and in translating detections into actionable estimates of importation and local transmission for public health decision-making. Here we develop a computational framework to evaluate strategies for respiratory pathogen detection that integrates an established clinical surveillance modality, intensive care unit (ICU) sampling, with an emerging environmental modality, aircraft wastewater (AWW) sampling. Detections are translated into risk via a multi-scale, stochastic global transmission model that combines international flight data with a detailed agent-based local transmission model. The resulting model-based estimates contrast the time to pathogen detection via AWW at airports with that in the community via realistic healthcare testing pathways. Using real-world data from England and Wales (EW), we find that employing AWW in EW airports can improve first detection times by 12.5-37.7 days for a range of epidemiological parameters under realistic healthcare testing scenarios and random aircraft sampling between 25 and 50%. In particular, for a SARS-CoV-2-like pathogen, we expect AWW to outperform ICU in first detection timing by 22.0-25.6 days, with ∼21.9-42.6 times fewer cases at their respective time of detection. While false detection remains a risk, we show that follow-up confirmatory testing can improve detection confidence substantially. Together our results demonstrate the potential utility of AWW surveillance and how it can reduce detection times and improve global health security.

## Introduction

Pathogen emergence and global dissemination have the potential to cause severe global economic, health and societal disruption (1). Increasing urbanisation, population growth and international travel mean that pathogens can spread faster and across greater distances from their location of origin (2). Further, climate variability and heterogeneous contact patterns reduce the predictability of pathogen emergence and increase transmission suitability (3, 4). Early pathogen detection and risk assessment in these changing contexts is therefore critical to the design and implementation of effective non-pharmaceutical interventions (NPIs) and the development of pharmaceutical countermeasures, whether to prevent further spread or to minimise disease burden (5, 6). Multiple concurrent efforts have been proposed to reduce the detection time of novel pathogens via coordinated and collaborative surveillance (7–9). What remains needed for improved risk assessment are frameworks that can quantify detection differences across different surveillance systems (e.g., global and national) and modalities (e.g., clinical vs. environmental) (10–12).

Genomic and metagenomic data enable rapid identification and characterisation of emerging infectious diseases (11, 13). Large-scale sequencing can reveal novel clades and their spread into globally connected regions, as shown for mpox in the Democratic Republic of the Congo (14). When paired with phylodynamic methods, these data can also reconstruct spatial dissemination, even when contact tracing is limited, informing targeted control measures and interventions (15, 16). In practice, however, most pathogen genomes still come from patient-focused surveillance. These samples can be biased toward severe infections and populations with better access to healthcare, and are often collected after transmission is already widespread (17, 18). While this clinical focus has strengths, including high-quality sequences and rich metadata, it can be complemented by wastewater sampling, which provides a population-level signal and captures community transmission. Integrating these information streams with global metapopulation transmission models informed by human mobility provides an opportunity to more rapidly translate detection into the risk assessments needed for effective responses.

Prior work has focused largely on optimising global or community/regional-level sentinel surveillance (7, 19, 20). Even in cases where frameworks for optimally integrating molecular and case data were developed, their evaluation was isolated to specific regions (21, 22). There remain open questions on how to integrate surveillance in the international travel network with local surveillance systems for the early detection of novel and climate-sensitive pathogens (23). Building on prior work (7, 24), we here develop a unified detection-to-risk framework that couples mobility-based importation with country-specific transmission and observation processes, including healthcare-seeking pathways informed by stakeholder input, clinical genomic testing designs in wastewater (WW, including aircraft and healthcare facility testing) and clinical-based surveillance. We seek to assess the time to detection of novel pathogens and provide estimates of the number of locally-generated secondary cases at the time of detection. We discuss how our work is scalable to other regions and could serve as a model for countries to design and evaluate their own pathogen detection frameworks in the context of global emergence and spread when resources are limited.

## Results

### Multiscale transmission model and surveillance system

We develop an integrated, multi-scale framework that couples global pathogen dissemination with within-EW testing and transmission (7) (Fig. 1A). At the global scale, locations are represented as metapopulations connected by international air travel, and pathogen introductions can occur anywhere in the network. Early spread is modelled as a multi-type branching process, with infection progression described by a Susceptible-Latent-Detectable-Recovered (SLDR) natural history (see Materials and Methods for details). Importations into EW via air travel then seed local transmission, which we simulate using an individual-based model (IBM) that captures movements from arrival airports using available commuting data ((25), Fig. 1B). Introduced pathogens can be detected via two detection routes. Firstly, there is detection in pooled AWW, which is collected daily, with detection dependent on lavatory usage, shedding probability, and proportion of aircrafts randomly selected for pooling. Secondly, detection in healthcare settings is determined by empirical healthcare utilisation pathway data in EW, where testing is distributed in ICUs (see conceptual overview in Fig. 1C). Importations are spatially distributed across EW according to the proportions of international air passenger traffic arrivals at EW airports, with onward local transmission incorporating commuting data (25). Infected individuals progress through a severity-stratified care pathway and, in our implementation of the adapted framework (24), only severe infections requiring ICU admission are assumed to be eligible for pathogen sequencing. We therefore model clinical detection exclusively via ICUs: 5% of infections progress to ICU, 10% of ICU patients will undergo metagenomic sequencing, and sequencing results are returned after three-days. The modelling framework is flexible and can be adopted for any setting for which appropriate data on the health care system, care and testing pathways are available.

**Figure 1:**
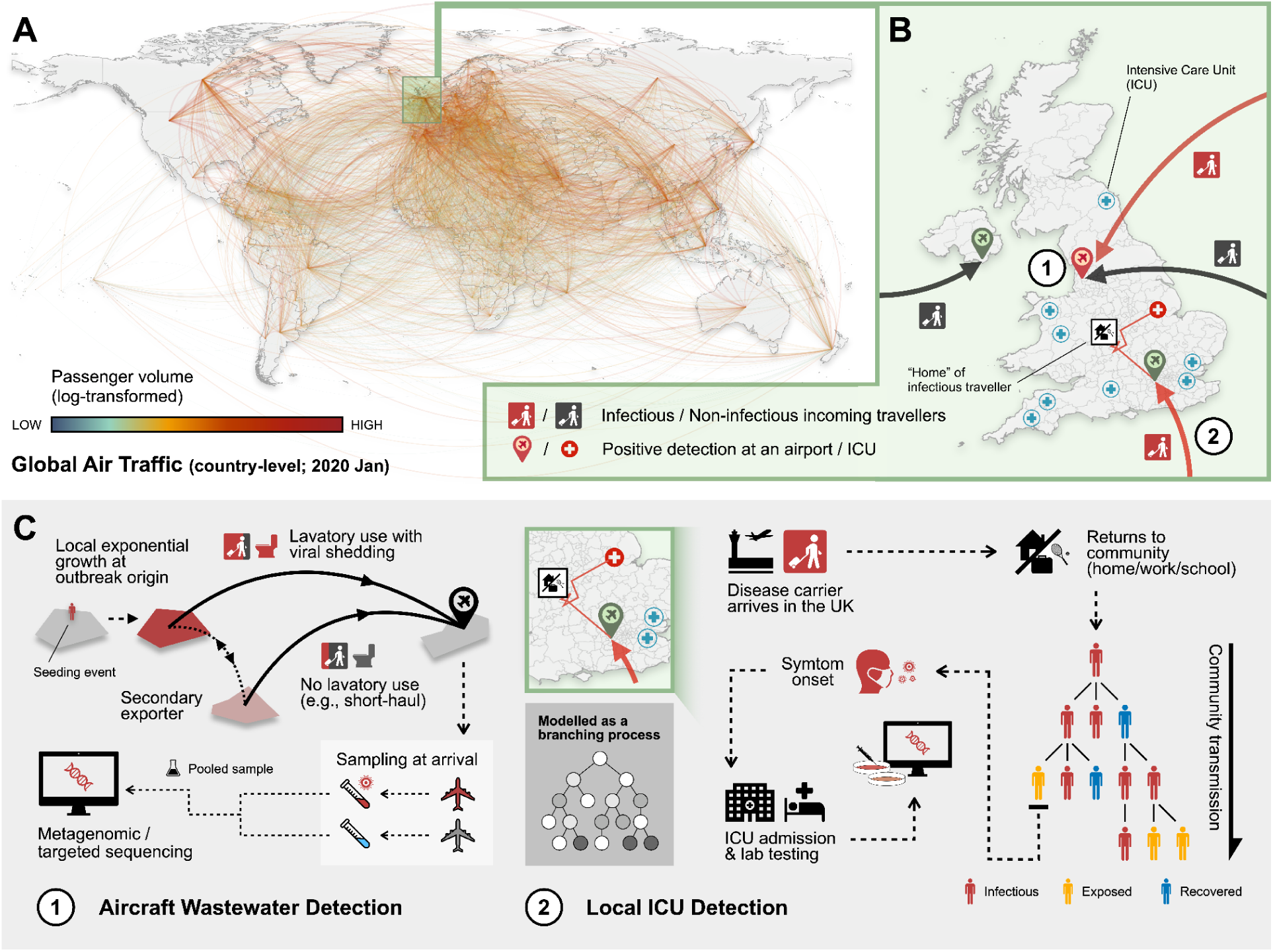
**A)** Global air traffic flows (country-level, January 2020) showing passenger volume (number of monthly passengers, log-transformed). **B)** Schematic of EW-based detection strategies: (1) aircraft wastewater surveillance at airports, and (2) healthcare detection of symptomatic cases in intensive care units (ICUs). Arrows show the arrival of infectious (red) and non-infectious (black) travellers into EW airports. Red lines in EW illustrate their spatial movement patterns, with markers indicating positive detections at airports (location pins) and ICUs (circled crosses). **C)** Methodology workflow illustrating: (**1**) Aircraft wastewater detection - disease carriers arrive via different routes (with or without detectable viral shedding in aircraft lavatories); pooled wastewater samples from arriving aircrafts undergo metagenomic or targeted sequencing. (**2**) Local ICU detection - following arrival of infected individuals in EW, transmission can occur via contacts in the community; in some cases infected individuals progress from mild to severe symptoms and ICU admission where laboratory testing is performed.

### Relationship between air travel, detection timing, and case burden

We use our simulation framework to consider a SARS-CoV-2-like virus, with a basic reproduction number of R_0_ = 2, and a 4-day generation time (infectious period = 2 days, latent period = 2.67 days) (26–28). We find that in the early phases of exponentially growing outbreaks, most of the first international importations originate from the location of outbreak origin (15, 29). This pattern holds regardless of the origin’s general connectivity or its relative position to the UK; even when an outbreak originates in the periphery of the global airline network, the high local incidence at the origin dominates early importation flux (30). We find that earlier arrival, defined as the time the first import arrives into EW, is predicted by a higher proportion of travellers from that country travelling to EW among all travellers, rather than by travel volume (Supplementary Fig. 4A).

Although the timing of the first importation into EW varies widely by outbreak origin (Fig. 2B), the subsequent lead-time (the time between the first import and detection) remains remarkably consistent. This is because the effective threshold of importations required to trigger an AWW detection is roughly equivalent across all scenarios, assuming a fixed per-traveller detection probability. Since we utilise consistent transmission and natural history parameters for the local EW model, the pathogen incidence grows at approximately the same exponential rate once introduced. Consequently, an increase in the time to first importation leads to a roughly equal increase in the time of AWW detection; the additional time required to move from the first arrival to a positive wastewater detection therefore varies little across origins, regardless of where the outbreak began (Fig. 2A).

**Figure 2:**
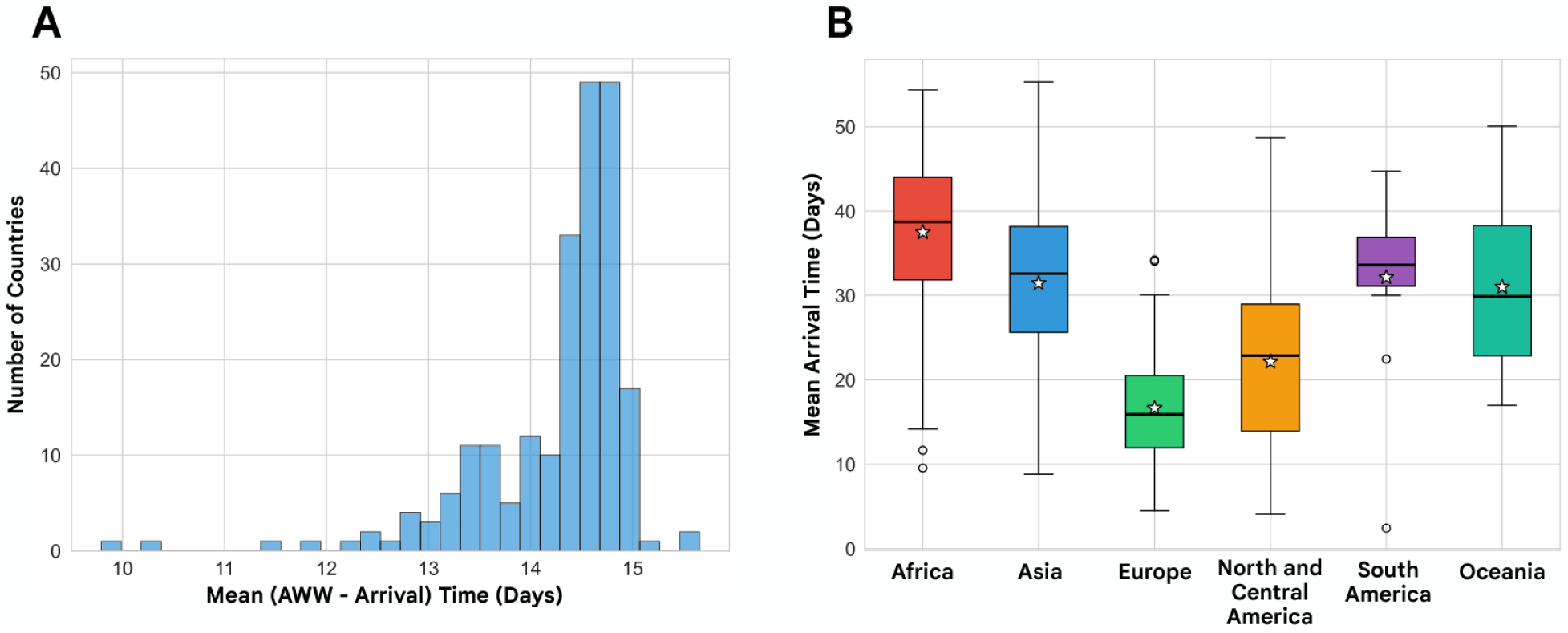
**A)** Histogram showing mean AWW detection lag time (i.e., difference in the mean AWW detection time and the mean arrival time) across all countries, with a mean of 14.3 days. Countries with the lowest differences have particularly high probability of travel to EW (outliers correspond to Gibraltar, Falkland Islands, Barbados and Iceland). **B)** Boxplot showing the distribution of the mean arrival times since the outbreak started, by continent of the outbreak origin (the horizontal line and the star in each box represents the median and mean of the country-level means respectively; both are aggregated across all countries in the same continent).

### Evaluating detection in healthcare vs. aircraft wastewater

The local model simulates pathogen transmission in EW following importation events generated by the global model (24). To ensure consistency, epidemiological parameters were kept the same between the global and local model. The ICU-based comparison reflects the constraints of the local model and provides a conservative, late-stage benchmark for clinical detection. Earlier healthcare encounters - such as urgent care or emergency department triage - would likely reduce the clinical detection lag relative to AWW surveillance, evaluating them remains outside the scope of this study. By integrating the importation dynamics from the global model (which provide wastewater detection time distributions) with the local ICU detection model, we directly compare first-detection timing under AWW surveillance with that under ICU-based clinical detection.

To capture stochastic variation in importation timing in the global model and local transmission dynamics, we ran 100 simulation replicates. For the specific case studies (Supplementary Figs. 6-17), we increased the number of replicates to 1000. Thus, observed differences in detection timing reflect the relative effectiveness of the underlying surveillance strategies, rather than consequences of the transmission model.

The correlation (r = 0.237, p < 0.001) between AWW and ICU detection times demonstrates that the detection difference is systematic, not an artifact of specific outbreak origins or importation patterns, highlighting that the variation in the differences in ICU and AWW detection times are small (Supplementary Fig. 4B). For a SARS-CoV-2 like virus, and 50% random sampling of aircrafts, the mean AWW detection was estimated at 25.6 (95% CI: 23.1-28.0) days earlier than ICU-based surveillance (Supplementary Fig. 3A). This timing gap has a large effect on secondary transmission: by the time of initial ICU detection, the cumulative number of local cases is expected to be 42.6 (95% CI: 41.5-43.6) times greater than at the earlier airport detection (Supplementary Fig. 3B). We present examples for outbreak origins in Paraguay, Ghana and Switzerland representing different positions in the airline network and find consistent differences in detection times and case differences (Fig. 3A).

**Figure 3:**
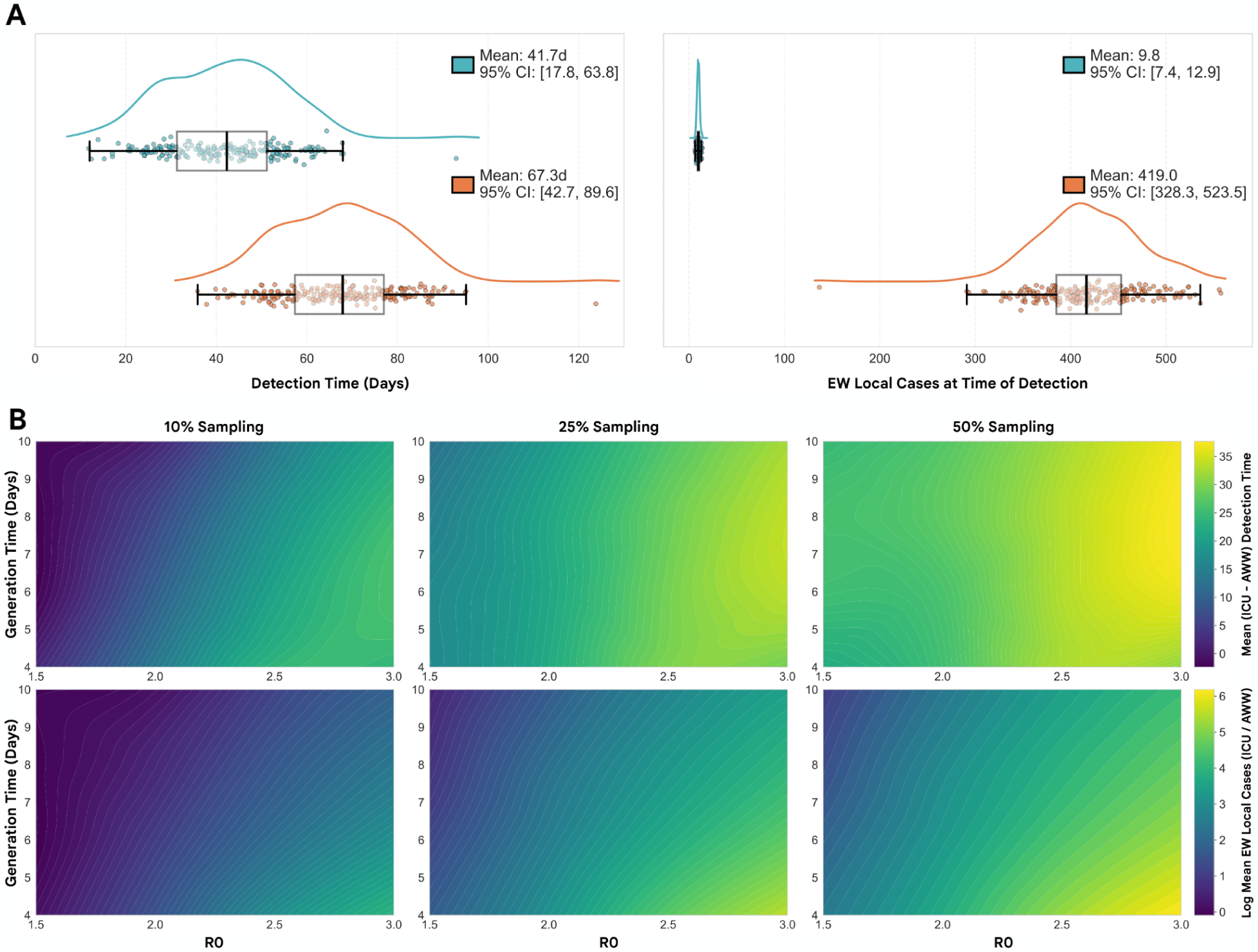
**A)** Country-level comparison of detection performance between surveillance methods. Left panels show estimated PDFs (derived via KDE) detection time distributions (days from outbreak start) for wastewater (blue) and ICU (orange) surveillance, with 100 samples per country as the candidate outbreak origin. Scatter points represent the means for each country as the outbreak origin, with box plots indicating median and interquartile range. Right panels display the EW local case burden at time of detection for the same outbreak origins, illustrating the trade-off between detection speed and epidemic size. **(B)** Heatmaps showing the estimated difference in mean detection time (top row) and difference in mean EW local cases at time of detection (bottom row) across parameter space; varying R₀ (x-axis), mean generation time (y-axis), and aircraft sampling rate (10%, 25%, 50% across columns), assuming an individual-level AWW detection probability of 16%, and ICU sampling rate of 10%. The filled contour levels represent interpolated surfaces of equal mean values (isoclines), generated using RBF interpolation on the mean simulation results, with 100 samples per country as the candidate outbreak origin. Lighter color indicates faster detection (top) and fewer local case at time of detection (bottom) for AWW surveillance.

We performed sensitivity analyses varying AWW sampling rate (10%, 25%, 50%), R_0_ (1.5-3.0), and mean generation times (4-10 days). We found that the difference in locally generated cases at the time of detection decreases as mean generation time increases (Fig. 3B, Supplementary Figs. 6-17). This is driven by the fact that a longer generation time results in a smaller growth rate, reducing both the rate of importation intensity into EW and the subsequent local transmission (Supplementary Fig. 1).

Conversely, for a fixed generation time, increasing R_0_increases the absolute difference in detection times between ICU and AWW surveillance. The most pronounced advantage for AWW - measured by the ratio of cumulative local cases at detection - occurred in ‘fast’ epidemic scenarios (high R_0_, low generation time), where rapid local growth causes ICU-based signals to lag significantly behind the actual infection burden.

### Spatio-temporal evolution of importation risk

Next, we sought to evaluate scenarios in which the origin location of a novel outbreak is unknown. We can construct a policy scenario in which an AWW detection is made and a question arises about from where the risk of importation is highest. In this case, we are further faced with uncertainty around the time of the initial outbreak, which influences where secondary outbreaks might have already started.

However, pathogen genomic data, which are now commonly collected as part of outbreak investigations, often provide hints about the timing and location of a novel spillover event or the timing of emergence of a novel pathogenic lineages/variants (e.g., (31, 32)). In the following, we evaluate how predicted importation risk changes over the course of an outbreak after a first detection is made but the origin location is unknown (Fig. 4A). Using the example of an initial positive detection on a flight from Nigeria to EW (and assuming, for simplicity, that the flight carries only travellers from the origin country and no transfer passengers), we find that the risk for importations to EW is expected to be heavily dependent on the time of emergence (Fig. 4C). Should the detection be early in the outbreak, then the risk of importations to EW is expected to be heavily concentrated from the outbreak origin (in this case assumed to be Nigeria, Fig. 4B). If the first initial positive occurs later, then the estimated number of plausible outbreak locations increases, and so more countries are predicted to be secondary exporters.

**Figure 4:**
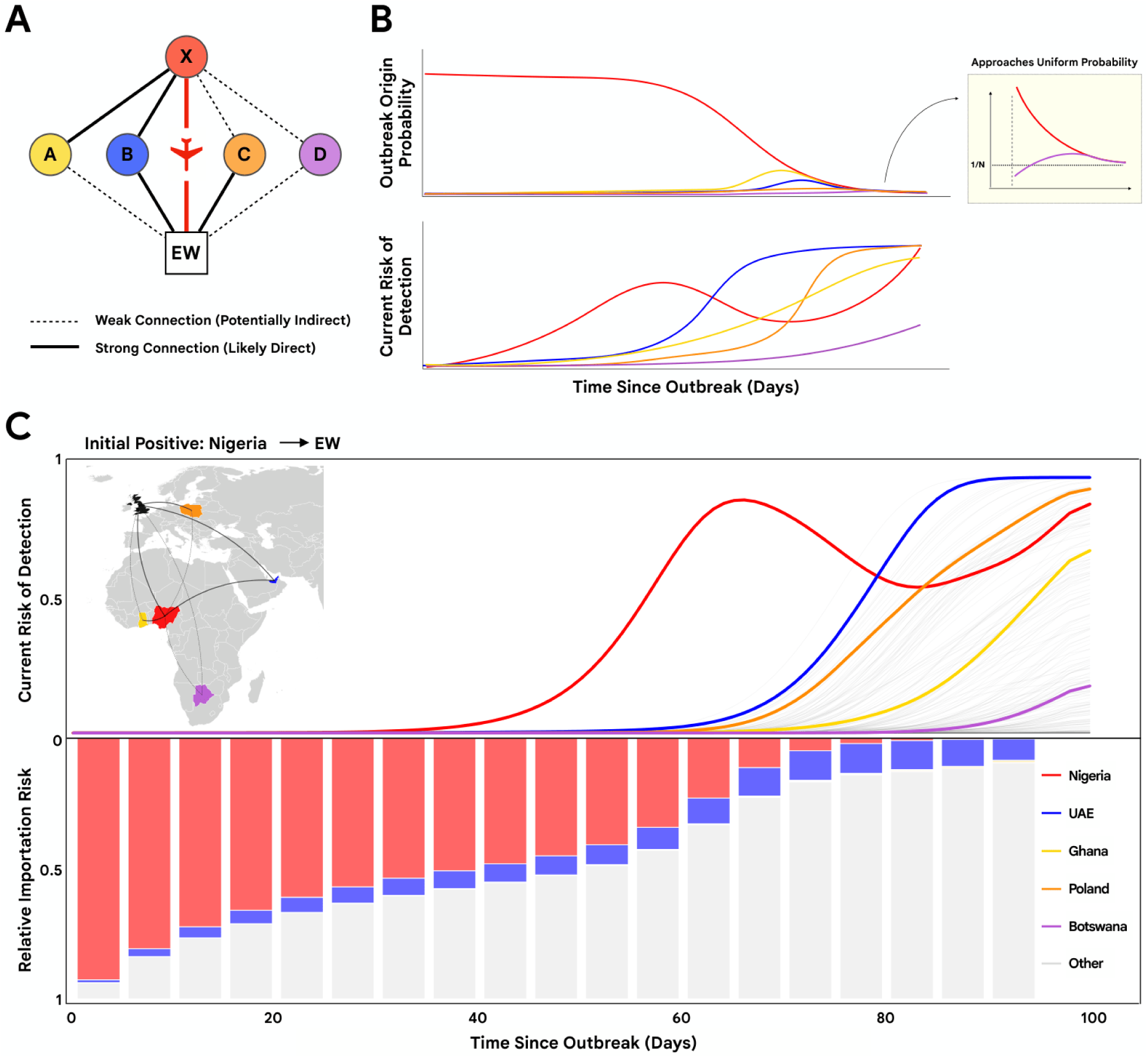
**A)** A schematic showing a hypothetical scenario in which through (AWW) surveillance, a positive detection on a flight into EW occurs, originating from some country X (red circle). The other four coloured circles represent countries in the global air traffic network categorised by their relative strength of connection to country X and EW: (A; yellow circle) countries with a high mobility flux (i.e. a strong connection) to/from country X but a low mobility flux (i.e. a weak connection, potentially through intermediate countries) to EW, (B; orange circle) countries with a strong connection to EW but a weak connection to country X, (C; blue circle) countries with a strong connection to both country X and EW, and (D; purple circle) countries with a weak connection to both country X and EW. **(B) Top:** Posterior probability of different countries being the outbreak origin given the initial positive X→EW detection, as a function of time elapsed since the start of the outbreak. In the limiting case where the detection is made as soon as the outbreak started, the most likely origin is X; with time, countries with a strong connection to both country X and EW become increasingly plausible (blue line), followed by countries with a strong connection only to country X (yellow line). Every country becomes equally likely to be the outbreak origin at large elapsed times, as indicated by the probability approaching 1/N (inset). **Bottom**: Conditional probability of a further detection of an infected arrival in EW from different countries, as a function of time elapsed since the start of the outbreak. The probability of a further detection generally increases with time regardless of the country of origin, as the overall infection prevalence increases across all countries. The estimated probability of a further X→EW detection however experiences a transient drop due to the rapid decrease in the probability of country X being the outbreak origin with time elapsed. **(C)** A specific case study where the initial country of detection is Nigeria. **Top**: Conditional probability of a further detection of an infected arrival in EW from different countries (equivalent to the bottom plot of panel B). Nigeria (red), United Arab Emirates (blue), Ghana (yellow), Poland (orange), and Botswana (purple) represent examples of the five different types of relative connection to the country from which a disease carrier is first detected and EW, with the same colour scheme as in (B); lines correspond to countries not highlighted are coloured grey. Bottom: Relative contribution of the risk of further importation, accounting for total import volume, from different countries, conditioned on different time elapsed since the start of the outbreak, at 5-day intervals.

The model plausibly recovers this changing risk landscape based on the structure of international passenger movements. Beyond identifying risk, this framework suggests that AWW can be resource-optimized by targeting the highest-risk flight routes. We find that sampling solely from the top three ranked candidate origins - identified by the expected time to first importation from each country, which accounts for the vast majority of initial importations - results in a similar mean detection delay of approximately 1 day compared to testing more widely (Supplementary Fig. 5). This highlights that flight-specific selection can significantly reduce sampling effort while maintaining early-warning sensitivity.

### Impact of test specificity

Detecting a pathogen soon after its emergence remains challenging due to imperfect detection, especially when early pathogen prevalence is low. We have assumed so far that a positive detection represents true pathogen presence (7). However, as discussed in (31), false-positives are a common problem at low prevalence. Although lab-based false-positive rates (FPRs) have been estimated to be about 0.8-4.0% (with targeted molecular approaches demonstrating higher specificity than metagenomic approaches (33)), even such low FPRs can lead to substantial numbers of false-positives compared to true-positives in low-prevalence settings. This has important implications for the utility of an early detection system, as false-positives could lead to unnecessary alarm and allocation of public health resources (30).

To address this, one possible approach is to implement a multi-stage testing protocol, where an initial positive detection is followed up with confirmatory testing. In this framework, a detection is only considered to be ‘valid’ if a certain number of consecutive AWW samples - each from different, successive aircraft arrivals from the same flight origin - return positive results. While this approach can reduce the probability of a false alarm, it also increases the time and resources required for detection, potentially delaying the identification of the presence of a developing outbreak. Here we examine this trade-off by exploring how the time-to-detection varies across different FPRs and number of consecutive positive tests required for confirmation (referred to as confirmatory test threshold (CTT) hereafter).

We estimate that a CTT of 1 (i.e. only a single positive test is enough to trigger a “valid” detection) is likely to be sufficient with FPR=0.8% (Fig. 5A, top row), with detection time relatively unaffected compared to a true system with no false positives. In contrast, when FPR=4%, detections occur considerably earlier compared to a scenario of FPR=0% (Fig. 5B, top row). While this may initially appear to be beneficial, many of these early detections are in fact false-positives, as indicated by the white bars in the histograms. Increasing the CTT to 2 or 3 (middle and bottom rows in Fig. 5A) reduces the false detection rate, as reflected by the shrinking size of the white bars. However, this comes at the cost of increasing time-to-true detection (as shown by the rightward shift of the coloured bars, indicating detections triggered by consecutive positive tests with at least one true positive). This delay arises mainly due to multiple consecutive positive tests required. A CTT of at least 2 is likely required to reduce the false detection rate to an acceptable level if the inherent FPR in the system is greater than 4% (Fig. 5D).

**Figure 5:**
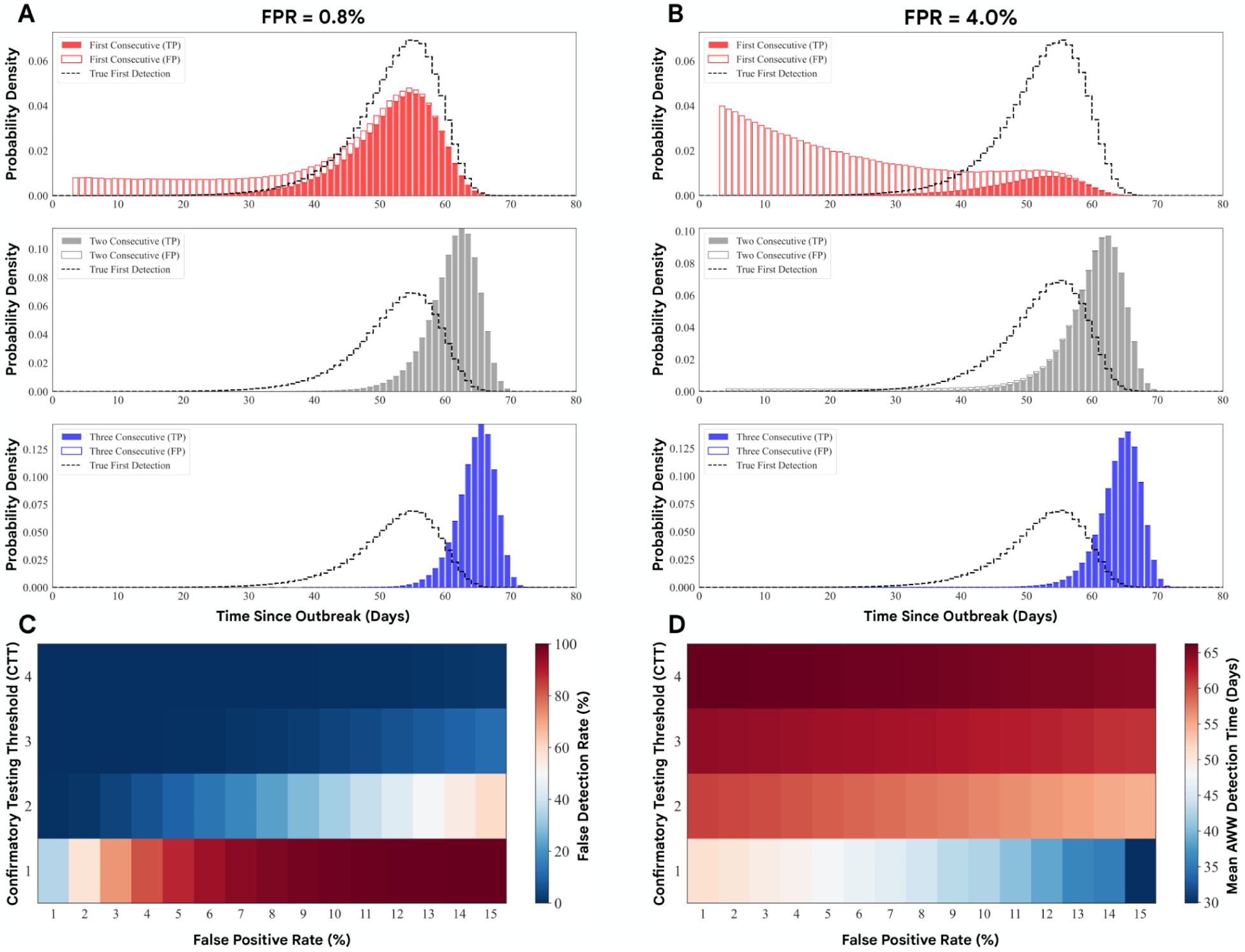
Consecutive testing resolves false positives at the cost of detection delay: Detection time distributions for **(A)** FPR = 0.8% FPR and **(B)** 4% FPR, corresponding with the expected lab-based ranges in (33), assuming an individual-level AWW detection probability of 16%. Each panel shows first positive (top), two consecutive (middle), and three consecutive (bottom) positives assuming a pooled testing strategy. True positives (colored bars) and false positives (white bars) demonstrate progressive increase in signal-to-noise ratio. With increasing FPR more consecutive positive results for confirmation are needed to avoid early false signals. Black dashed line: true first detection distribution (FPR=0%). **(C-D)** Heatmaps show the true detection rate, and the (true) mean detection time (the time to the first positive signal, assuming at least one of consecutive positives is true) across FPR levels and consecutive test requirements.

As the false positive rate (FPR) increases, the mean time to true detection decreases, particularly at lower CTTs (Fig. 5D). This does not reflect an improved signal-to-noise balance, nor does it represent a meaningful trade-off that can be optimised in practice, since the FPR is not a tunable parameter but rather an intrinsic property of the testing setup. Instead, the effect arises from how the decision rule interacts with background noise. A higher FPR increases the overall frequency of detection triggers, especially early false-positive signals, as shown in Fig. 5A (top row). For a fixed CTT, more frequent triggers increase the probability that the required sequence of consecutive positives is achieved earlier in time.

Consequently, the outbreak may be formally determined sooner dependent on a policymaker’s chosen CTT, even though the underlying epidemiological signal has not strengthened. Importantly, the magnitude of this apparent acceleration is modest and remains small compared to the delays introduced by increasing the CTT. Thus, while higher FPR can shift the timing of outbreak determination earlier, it does so by increasing noise and false alarms, and does not represent a practically desirable or controllable pathway to earlier detection.

## Discussion

In this study we introduce a multi-scale framework for evaluating pathogen detection strategies that integrates aircraft wastewater surveillance with healthcare-based genomic monitoring in the United Kingdom. In our idealised setting, the results indicate that AWW surveillance detects newly emerging respiratory pathogens earlier than surveillance focused in ICUs, with detections occurring 15-49 days earlier. However, our approach considers only respiratory pathogens and will need to be tested and benchmarked on other pathogens with varying levels of severity and test sensitivity. Theoretically, an earlier detection results in substantially fewer locally generated cases at the time of detection, providing additional time for public health interventions. Furthermore, our analysis demonstrates that the relationship among air-travel connectivity, detection timing, and EW case burden is complex.

Well-connected regions exhibit higher case counts at detection despite faster detection times, likely due to the combination of large numbers of importations and fast-growing epidemics in the source location.

The operational viability of AWW surveillance is fundamentally tied to its ability to distinguish true signals from background noise. As our results highlight, inherent false positive signals in early epidemic scenarios, as well as non-epidemic scenarios, can lead to misallocated public health resources and the potential for unnecessary operational interventions. Practical implementation faces significant empirical challenges, including variable detection sensitivity due to viral shedding patterns, dilution effects, and potential cross-contamination between flights if wastewater tanks are not thoroughly cleaned (34–38).

While we show that consecutive testing of a sequence of arriving aircrafts from the same origin acts as a powerful filter - in which a second or third positive can provide the confidence necessary to trigger a response - the effectiveness of this strategy depends on rapid laboratory turnaround times to maintain a temporal advantage over clinical detection. Initial implementations with airport authorities, and public health agencies, will be essential for operational refinement and for validating that the theoretical times observed in the model outputs are achievable in the presence of real-world logistical constraints.

Early pathogen detection provides benefits across diverse infectious disease threats. Wastewater surveillance networks can improve situational awareness by reducing detection time and addressing key limitations of symptom-based passenger screening (39, 40). Respiratory viruses such as influenza and SARS-CoV-2, vector-borne diseases including Zika and dengue, hemorrhagic fevers like Lassa and Ebola, Mpox and emerging zoonotic pathogens continue to pose threats to global health security (41, 42).

Extending our modelling framework to other pathogens is an important next step. Sensitivity analyses across different epidemiological parameters and detection probabilities (Supplementary Tables 1-16) demonstrate that, while the absolute detection gap between AWW and ICU narrows for pathogens with slower growth rates, the relative utility of AWW as an early warning tool remains robust. This suggests that for many emerging threats, geographic connectivity, rather than specific pathogen biology, is a primary driver of detection dynamics. However, this generalisability will be contingent on the underlying transmission mode; our framework assumes pathogens spread primarily through community contact.

Pathogens that spread through alternate pathways, such as sexual transmission networks, would likely exhibit distinct spatiotemporal and network dynamics that require different modelling approaches.

Furthermore, the relative advantage of one modality often depends on the objective. While AWW can offer broad community surveillance for a set of pathogens, e.g. those prioritised by organisations including the Coalition of Epidemic Preparedness and Innovation (CEPI) and the World Health Organization (WHO), clinical surveillance in ICUs targets severe cases.

The effectiveness of surveillance strategies depends heavily on context. Knowledge of the outbreak origin enables substantially faster detection through targeted sampling of high-risk routes, as shown in our case study. This finding highlights the importance of global surveillance coordination and rapid information sharing. When outbreak locations are known, countries can adapt surveillance strategies to maximise detection sensitivity for high-risk importation pathways.

Our framework extends beyond EW. Any country with detailed data on healthcare-seeking behaviour, clinical care pathways, and within-country population movement can adapt this modelling approach to design and evaluate country-specific surveillance strategies. The key requirement is obtaining realistic local transmission dynamics and healthcare system structure. This is essential because detection time depends on both pathogen arrival and subsequent community spread patterns, including the timing of infected individuals’ entry into surveillance systems. Accurate models of internal spread and healthcare seeking substantially improve assessment quality and enable more precise resource allocation.

### Limitations and Future Work

Our analysis has several important limitations. The model treats air travel as an independent process for individuals, neglecting clusters of individuals - such as groups or families travelling together or within plane transmission, which may affect early-stage outbreak dynamics. We used multitype branching process approximations that ignore saturation effects arising from finite population sizes; although these effects are minor during early outbreak phases, they may become significant when analysing surveillance performance in large epidemics or endemic situations. As such, our approach is valid only for early epidemic scenarios. Future work should focus on incorporating more diverse models of superspreading, demographic stochasticity, and other socio-biological factors known to influence pathogen transmission (43).

Our baseline ICU-based detection times should be viewed as conservative. In reality, detection time is highly pathogen-specific; any agent with higher clinical severity or a shorter window for seeking healthcare would likely yield faster detection than our respiratory model suggests. Additionally, our ICU model represents an early-stage infrastructure with a limited number of sites. As these systems evolve - for example, the UK’s proposed expansion of genomic surveillance infrastructure through the mSCAPE program - the comparative advantage of AWW over clinical detection may diminish (44).

Further, it is important to note that the earliness of AWW detection is not an absolute rule; during the COVID-19 pandemic, intensive community sampling and active case-finding enabled the UK to detect initial cases rapidly, outpacing what would be expected from passive ICU-based monitoring alone. Our local model assumes consistent healthcare-seeking behaviour and diagnostic testing rates across populations and over time, which may not hold during rapidly evolving outbreaks when healthcare systems face capacity constraints or behavioural factors influence care-seeking patterns (45).

Additionally, we have not incorporated heterogeneity in airline network structure at the flight level due to data limitations; instead, we rely on passenger-origin country data. Access to detailed flight-specific connectivity data would enable aircraft-specific sampling strategies, allowing the transformation of model outputs from passenger-origin level to the specific flights arriving into EW airports. In the first instance, with aircraft-specific data, we will transform our results provided in Figure 4 to now consider which flight routes to test in the UK, as the initial positive that we receive is from a flight into the UK. The combination of flight and passenger origin level data means we can combine the notion of countries with local outbreaks, and the composition of passengers on a flight to create a synchronised modelling and testing strategy.

The feasibility of metagenomic surveillance in aircraft wastewater remains a significant challenge. Our modelled individual detection probability, p_det_, reflects several underlying processes: the probability of lavatory use (which varies with flight duration), the pathogen-specific probability of viral shedding, and the sensitivity of metagenomic assays. Further, the type of metagenomic assay has a large influence on the efficacy. Untargeted metagenomic sequencing offers the advantage of agnostic detection for novel pathogens, but typically at the cost of a higher limit of detection (LoD), due to the background of host and commensal genetic material. On the other hand, targeted metagenomic approaches, which are used for a known pathogen, can significantly increase the effective detection probability of the wastewater test.

Following previous work, (7), we adopt a baseline constant value of p_det_ = 0.16, to represent a moderate detection regime. Crucially, our model evaluates detection at the aircraft level; if n infectious individuals are on a flight, the probability of at least one shedding event (which we classify as a detection) is 1- (1 - p_det_)^n^. However, the effective detection probability in practice could be higher or lower than this aggregate assumption, because it depends on pathogen and context specific factors that we do not model. These include: the flight occupancy (which contributes to the dilution of the signal by unfinfected passengers), distribution and timing of shedding across individuals, the extent to which biological material enters wastewater via faeces/urine versus other routes (e.g. respiratory secretions and hygiene behaviours), sampling and processing effects (e.g. mixing, degradation, inhibitors, recovery efficiency) and the performance characteristics of the assay. We therefore interpret false-negative risk as conditional on these operational and biological details.

Future research must also quantify the economic costs and cost-effectiveness of AWW surveillance relative to expanded clinical monitoring. The operational viability of such a system depends on the ’cost per day of lead-time gained,’ including the direct costs of high-throughput sequencing and the indirect socioeconomic costs of responding to false alarms. Requiring a higher confidence threshold of the truth value of a positive signal can reduce these indirect costs, but at the expense of lead time, revealing an inherent trade-off between cost mitigation and early detection. Effective early detection systems would require collaboration across private, public, and academic sectors (38). Airport and airline operators might control access to wastewater sampling infrastructure, public health agencies define surveillance priorities and response protocols, commercial laboratories provide testing capacity, and academic institutions contribute expertise and analysis capabilities (46). As discussed in earlier sections, the minimal performance degradation from only sampling the top 3 origin locations provides a simplistic understanding of how we might optimise the selection of flights.

Although further validation in real-world settings is required, our computational analysis indicates that aircraft wastewater surveillance can supplement traditional healthcare-based detection. Our work provides a methodological foundation for designing surveillance strategies that are both scientifically sound and operationally feasible, supporting the global goal of preventing the next pandemic through early detection and rapid response.

## Acknowledgements

Data and code availability:

## Author contributions

B.K.R., J.L.-H.T., M.U.G.K., A.V., K.O.D., E.V., M.C. conceived the study.

B.K.R., J.L.-H.T., K.O.D., E.V. G.S., J.D., M.U.G.K., A.V. designed the methodology. B.K.R., J.L.-H.T.,

K.O.D., E.V. G.S., J.D. analysed the data and ran simulations. B.K.R., J.L.-H.T., M.U.G.K. wrote the manuscript. All authors commented on and edited the manuscript.

## Materials & Methods

### Probability generating function framework for the global transmission model

We describe early international spread, importation, and detection using a probability-generating-function (PGF) methodology introduced in the aircraft-wastewater surveillance framework, developed in (7, 47). The PGF formulation provides an analytic representation of the stochastic, mobility-structured branching process underlying the Global Epidemic and Mobility (GLEAM) model, which couples metapopulation transmission with global human mobility (48). In particular, we build on the GLEAM modelling foundations and applications described in the context of global dissemination and early cryptic spread analyses, and on recent multi-scale extensions, that link global dissemination to country-level dynamics (49, 50). Here we use a formulation defined at the geographical resolution of individual countries, rather than the urban area level used in (7). We denote this as the *global model* throughout. The PGF provides an analytical framework for representing the stochastic dynamics governing the initial phase of an outbreak, enabling evaluation of detection probabilities across all potential sources without the computational cost of large simulation ensembles. At each time step t, the system is represented by a state vector s = [s_1_,…, s_N_]^T^, which records the number of individuals in different epidemiological stages, age groups, and subpopulations, as well as cumulative exported cases and detections. The probability distribution for this state is encoded through

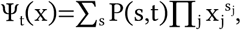

The stochastic elements of the model follow directly from GLEAM. Transmission arises from inhomogeneous Poisson processes: during each short interval Δt, the number of new infections is Poisson-distributed with a mean determined by the local force of infection, which varies over time and across subpopulations. Infection progression is described by a Susceptible-Latent-Detectable-Recovered (SLDR), where latent individuals are allowed to onward transmit but remain undetectable in AWW, infected individuals are capable of transmission and detected in AWW, and post-infectious individuals can no longer transmit, but are still capable of being detected in AWW. This mechanism is treated as a set of binomial transitions, reflecting the residence-time distributions implied by the compartmental scheme.

Daily mobility is modelled using multinomial draws based on travel probabilities between subpopulations, and is not conditioned on individuals’ disease or symptom status. As a result, symptomatic or severely ill individuals are assumed to travel at the same rates as others, which may overestimate early importation for more severe pathogens. Detectable travellers are identified at sentinel airports via Bernoulli sampling along flight paths. Together, these ingredients give a multitype branching-process description of the early outbreak phase, consistent with the formulation in the SI of (7).

Quantities such as the distribution of cumulative importations can be extracted directly from the PGF. To do this, we apply a mapping A(x) that replaces the dummy variable with x for components that record importations and with 1 elsewhere. The resulting marginal PGF ζ_t_(x)=Ψ_t_[A(x)] gives probabilities such as P(d_t_=0)=ζ_t_(0), from which the time of first detection follows

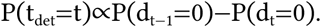

Thus, our framework yields the full distribution of first-detection times.

Following the global model, each day is divided into a reaction phase and a mobility phase. The reaction phase (modelling transmission and disease progression) is described by a vector of PGFs r(x), applied n_r_ = 12 times per day to account for sub-daily changes; mobility and detection are encoded in a second vector M(x). Combining them gives the daily update R(x) = r^(nr)^(x), F(x) = M(R(x)), and the PGF evolves through

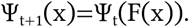

Repeated composition from the initial condition Ψ_0_, which we fix to be 10 exposed and 10 infectious individuals in the outbreak origin, yields the full time evolution of importations, onward transmission and detection. This framework allows us to evaluate early-detection timelines across the global mobility network while retaining the stochastic features that are important during the early phase of an outbreak.

### Wastewater Detection Probability

When there are multiple flights from the same origin per day, we can model the surveillance system as testing M randomly selected flights out of K total flights. Under the assumption that infectious individuals are randomly distributed across flights and detection events are independent, the probability of detecting at least one positive flight is 1 - (1 - p_sam_·p_det_)^N^, where N is the total number of infectious individuals across all flights and p_sam_ = M/K is the fraction of flights tested. This equivalence between discrete flight sampling and aggregate detection holds specifically for the binomial detection model we employ throughout. In the case of a different flight-level detection model, the pooling and flight-level protocols don’t align. In this way, we evaluate a range of p_sam_ values (10%, 25%, 50%, 100%), which is consistent regardless of whether we pool all wastewater samples into one and obtain a positive detection, or take samples of aircrafts individually and identify at least one positive aircraft.

### ICU-Detection Model

The local transmission model simulates pathogen detection through ICU-based metagenomic surveillance following importation events estimated from the global model. The model is implemented in Julia and source code is available at https://github.com/emvolz/NBPMscape (manuscript in preparation (24)). A key analytical challenge is that we are estimating extreme value statistics for the importation data - specifically, the distribution of first detection times across stochastic epidemic realisations. This distribution is complex and lacks closed-form theoretical results; it remains understudied in standard sampling theory and necessitates simulation-based inference.

The NBPMscape model is a multi-type stochastic branching process in continuous time, implemented using the JumpProcesses.jl Julia library (51). The software architecture is designed to be modular and easily configured for different epidemic scenarios, allowing users to adjust epidemiological parameters, surveillance methods, and population structure as needed. Multiple simulation replicates (typically 100–1,000) are run to characterise the distribution of detection times and locally-generated cases under each scenario.

We adapted this model to simulate local transmission within EW following importation events, and denote it the *local model*. The model is an individual-based branching process, in which onward transmission can occur following modelled importations. To ensure consistency between the global and local models, we aligned the epidemiological parameters.

#### Model Components

1. Importation dynamics: Importation events are not generated stochastically within the local model; rather they are produced by the global model. Importations are then distributed spatially across EW regions according to the proportion of international passenger traffic each airport receives, based on 2024 data from the UK Civil Aviation Authority (52). Once infected travellers arrive, they move around according to commuting patterns we derived from 2021 UK Census origin-destination data (53), which captures how people travel between home and work locations.
2. Disease natural history: Latent periods, infectious period duration, and generation time distributions follow gamma distributions calibrated to COVID-19-like transmission dynamics.
3. Age structure: The model incorporates age-dependent disease severity and age-dependent travel patterns. The age distribution of imported infections is realistically modelled based on international traveller demographics.
4. Geographic structure. Spatial heterogeneity is captured through commuting patterns derived from UK Census origin-destination survey data, determining how individuals move between residential and work locations at the ITL2 level.
5. ICU sampling probability. The probability of an infection being detected combines two components: (i) the probability of having a severity profile leading to ICU admission, and (ii) the probability that the admitting ICU falls within the surveillance catchment areas, or optionally a tuneable uniform-random sampling probability for all ICU cases.
6. Contact heterogeneity. Transmission occurs through contacts in three settings: household, workplace/school, and community. Contact patterns are age-dependent and parameterised using POLYMOD survey data. POLYMOD UK survey data (54) characterised workplace/school and casual contacts, and household contacts. Household contacts have the highest transmission probability (baseline = 1.0), while workplace and casual contacts are scaled down to 50% of this baseline rate, reflecting shorter contact duration and lower transmission risk (55).
7. Alternative surveillance modules. An optional module captures detection through general practitioner consultations, as well as non-ICU hospital admissions, where detection probability depends on the individual’s severity profile.
8. Temporal dynamics: Contact patterns vary temporally based on commuting behaviour. Individuals commute to work or school locations where they accumulate workplace/school contacts. Day-of-week effects modulate contact rates, with higher contact frequencies on weekdays.

The local transmission model does not accept R₀ directly as a parameter, but instead uses an infectivity factor that modulates the per-contact transmission probability. To achieve our target R_0_ values which we explored in sensitivity analyses, we calibrated this infectivity parameter by simulating individual infections across household (F), workplace (G), and random (H) contact networks, counting secondary infections by contact type, constructing a next-generation matrix, and computing its dominant eigenvalue to determine the resulting R_0_. We determined that the default NBPMscape infectivity value of 1.25 produces R₀ ≈ 2.03, validated across 100000 MC samples. Using this baseline, we employed proportional scaling to set infectivity values for other target R₀ values.

For the disease parameters, we chose values that match COVID-19-like transmission. The latent period was varied to achieve a mean generation time of 4.0 days, fixing the mean infectious period at 2.67 days (8/3 days) as a biological constant. We adapted the local model, which by design integrates gamma distributions for latent and infectious periods, to output near-deterministic distributions by having large shape parameter (we arbitrarily set shape = 1000). By fixing the infectious period when varying the generation time, this approach ensures that R₀ varies only through changes in transmission probability per contact, while the duration of infectiousness remains constant across all scenarios, matching the compartmental structure assumptions of the global metapopulation model.

Disease severity, disaggregated by age, determines how individuals progress through the healthcare system. WHO estimates were used to classify infections (56) 60% develop mild or asymptomatic infection, 35% moderate infection, and 5% severe infection (57). Those with moderate or severe infection visit a GP at a rate of 1/3 per day. Severe cases can be admitted to hospital (rate of 1/4 per day) and then to ICU (rate of 1/5 per day), following progression rates adapted from (57). Once patients are hospitalized or admitted to ICU, their transmission probability drops by 75% (multiplied by 0.25) due to infection control measures like isolation and protective equipment.

For ICU-based surveillance, we implemented a sampling strategy where 10% of individuals admitted to any ICU undergo metagenomic sequencing, reflecting realistic operational constraints observed in pilot implementations (58), although this will likely be increased to approximately 30% in the coming years. After a sample is collected, we impose a 3-day turnaround time for sequencing and bioinformatics analysis before a novel pathogen would be identified. While optimised research workflows can achieve same-day results, typical clinical metagenomic sequencing turnaround times range from 24-72 hours (59).

### Integrating the global and local models

The global importation model provides time-varying importation distributions, which we discretise for use in the local model. In particular, we are able to extract time series of the total number of travellers into EW by mechanistic compartment, i.e. latent, infectious and post-infectious individuals. Following (7), infectious and post-infectious individuals can be detected in wastewater, but we assume that latent individuals move into EW undetected. When seeding imports into the local model, we restrict to latent and infectious individuals; post-infectious individuals are no longer capable of infecting others.

In particular, as the global model is not individual-based, we assume each latent individual is halfway through the latent stage, and similarly infectious individuals are halfway through their infectious stage, upon arrival into EW. Given the underlying exponential growth, we would likely expect more individuals to be at the start, rather than the end, of each stage; as such, we are potentially underestimating the average wait-time until latent individuals can start transmitting in EW, but on the other hand also underestimate the number of secondary cases existing infectious individuals may cause. These effects partially offset one another, however as we increase the latent period in sensitivity analyses, we expect that the reduced latent duration is more prominent; therefore resulting in a slightly earlier ICU detection time than present.

Stochasticity enters the local model framework in two distinct ways. First, local transmission dynamics are governed by the continuous-time, multi-type stochastic branching process presented above. Second, importation seeding is treated stochastically; we generate discrete arrival counts by drawing Poisson samples from the aggregate of latent and infectious imports at each time step. These two sources of stochasticity ensure that each simulation replicate reflects both varied introduction events and divergent transmission trajectories.

### Targeted Route Sampling for Confirmatory Detection

We adapted the source identification methodology from (3) to address the optimal validation strategy for EW-based aircraft wastewater surveillance. Given an initial positive AWW detection on route X → EW at time t, we seek to identify what other routes, V → EW, should be tested to confirm the initial positive signal; and in turn, identify which other regions are of high risk.

#### Bayesian Source Identification

When route X → EW tests positive at time t, we infer the posterior probability of each potential outbreak origin using Bayes’ theorem:

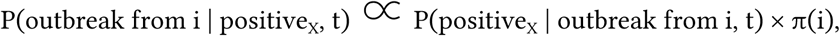

where π(i) is a uniform prior over all countries i, with non-zero import flows to X, and

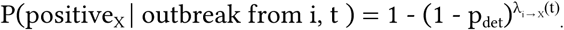

Here, λ_i → X_(t) represents the expected number of imports arriving on route X from outbreak source i at time t, given individual-level detection probability p_det_.

The posterior weights are:

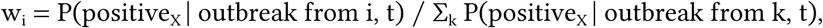

normalised such that they sum to 1; forming a posterior probability distribution for Bayesian inference.

#### Validation Route Selection

For each candidate confirmatory route V → EW, we calculate the expected confirmation probability by marginalising over all plausible outbreak sources:

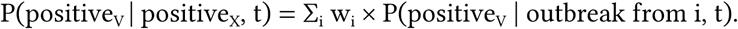

This represents “Given the outbreak scenarios that could plausibly explain the positive on X → EW, there’s a P(positive_V_ | positive_X_, t) chance that V → EW would also test positive, at time t, if sampled”

To quantify the relative infection import burden from each source country, we calculate the proportion of total expected infections arriving at EW from each route, aggregated across all possible outbreak origins:

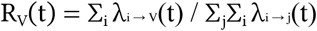

Where λi → V(t) represents expected imports from outbreak origin i to validation route V at time t. This metric is independent of detection probability as it reflects the volume of potentially infected travellers.

Moreso, we can identify the maximal validation strategy, which identifies the route that maximises this expected confirmation probability:

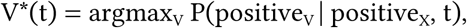

where V represents the set of all candidate validation routes.

### Test Specificity

Throughout this study, we have been considering the ‘time to first positive detection’, however this is an unobserved quantity. In reality, when we obtain the first positive signal, this contains the true positive signal, but also some underlying noise. The reliability of wastewater surveillance is defined by the probability that a positive detection sequence reflects a genuine pathogen importation event rather than an accumulation of false-positives. We evaluate this through two primary metrics: *signal reliability* (the proportion of alarms that are true positives) and the *operational delay*.

#### Simulation of Detection Chains

We utilise a Monte Carlo approach to simulate independent outbreak realisations for each country. From the global model, we generate the true importation trajectory μ(t) = [μ_0_, μ_1_, …, μ_T_] - representing the expected number of daily importations into EW. On any given day with a mean importation rate μ_j_, the test outcome is a function of both the biological detection process and the test’s inherent specificity. We define the model parameters as

● Specificity (1 - α): where α = False-positive rate
● Sensitivity (1 - β): where β = False-negative rate

For a day with n imports, a *positive* result can occur in two ways.

1. True positive: Pathogenic material is captured in the sample, with probability 1-(1-p)^n^, and the test correctly returns a positive result, with probability 1-β.
2. False-positive: No pathogenic material is captured in the sample, with probability (1-p)^n^, but the test incorrectly returns a positive result, with probability α.

This gives the daily probability of a positive detection, given n imports as:

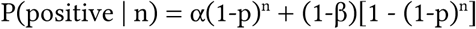

In our analysis, we assume the assay is highly sensitive once a pathogen is present in the sample, setting β=0. This implies that false-negatives in our system are driven primarily by the stochasticity of the sampling process (the ’per-import detection probability’ p) rather than assay failure. Under this assumption, the second term of the negative probability drops out.

To resolve the ambiguity of initial signals, we model a sequential verification protocol where an actionable alert is only triggered following N consecutive positive results. We consider two temporal parameters: *test frequency*, representing the frequency of the routine aircraft samples (eg. daily, once every 3 days), and *retest delay*, which is the turnaround time from sample collection to result availability (set to K=3 days, following (59)).

A detection sequence of length N is initiated when routine surveillance first detects a positive signal at time T₀ (sample collection day). Subsequent samples are collected at intervals of f days: T₀, T₀+f, T₀+2f, …, T₀+(N-1)f. Results for each sample arrive K days after collection. The chain is successfully completed only if all N samples return positive results. If any sample in the sequence is negative, the chain is broken, and the system resets to monitor for a new initial signal.

We define the *signal reliability* as the empirical true positive rate (TPR) of a fully completed sequence

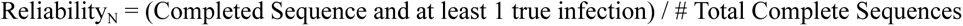

A sequence is classified as a “true positive” if at least one sample in the chain corresponded to an underlying true positive signal (i.e. was sampled on a day with n > 0 imports).

The recorded confirmation day is when the final N^th^ test result becomes available:

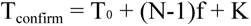

where T₀ is the initial detection day (sample collection), (N-1)f is the time to collect all N samples, and is the laboratory delay for the final result.

## Data Availability

We collected the monthly global flight and passenger data from January 2020 from the International Air Transportation Association (IATA) database, (https://www.iata.org/) which is accessible at https://zenodo.org/records/7472836. The IATA database contains information on flights between 4,418 commercial airports worldwide and can be expected to have a 100% coverage of the global airline market (60). To simulate the spatiotemporal dynamics of transmission at a global level, we aggregated the airport-level flight data into country-level origin–destination passenger flows. To minimise the number of assumptions and parameters, we evenly distributed the monthly passengers at daily intervals. Population estimates were taken from the United Nations Department of Economic and Social Affairs, Population Division (https://population.un.org/wpp/), specifically the 2019 revision for the year 2020; such that it aligns with the global flight dataset.

## Code Availability

The code associated with the work undertaken in this paper is publicly available at https://github.com/breddyk/AWW_and_ICU. The code associated with global model is publicly available at https://github.com/mobs-lab/pgfgleam, and the code associated with the England and Wales epidemiological model is available at https://github.com/emvolz/NBPMscape

## References

1. World Bank Group, “Chapter 1. The economic impacts of the COVID-19 crisis” (World Bank Group, 2022).

2. R. E. Baker, et al., Infectious disease in an era of global change. Nat. Rev. Microbiol. 20, 193–205 (2022).

3. R. de Abreu, et al., The Role of Climate Change in the Expansion of Dengue. Epidemiology (2025).

4. A. R. Kaye, et al., The impact of natural climate variability on the global distribution of Aedes aegypti: a mathematical modelling study. Lancet Planet. Health 8, e1079–e1087 (2024).

5. UKHSA Advisory Board: preparedness for infectious disease threats. GOV.UK. Available at: https://www.gov.uk/government/publications/ukhsa-board-meeting-papers-january-2023/ukhsa-advisory-board-preparedness-for-infectious-disease-threats [Accessed 24 August 2025].

6. Module 2, 2A, 2B, 2C Report - Core decision-making and political governance UK Covid-19 Inquiry Archives. UK Covid-19 Inquiry (2025). Available at: https://covid19.public-inquiry.uk/documents/module-2-full-report/ [Accessed 24 November 2025].

7. G. St-Onge, et al., Pandemic monitoring with global aircraft-based wastewater surveillance networks. Nat. Med. 31, 788–796 (2025).

8. E. Aßmann, et al., Augmentation of wastewater-based epidemiology with machine learning to support global health surveillance. *Nat*. Water 3, 753–763 (2025).

9. Z. Du, et al., Optimizing COVID-19 surveillance using historical electronic health records of influenza infections. PNAS Nexus 1, gac038 (2022).

10. J. L. Gardy, N. J. Loman, Towards a genomics-informed, real-time, global pathogen surveillance system. Nat. Rev. Genet. 19, 9–20 (2018).

11. N. C. Grassly, A. G. Shaw, M. Owusu, Global wastewater surveillance for pathogens with pandemic potential: opportunities and challenges. Lancet Microbe 6, 100939 (2025).

12. R. Hill, et al., Realising a global One Health disease surveillance approach: insights from wastewater and beyond. Nat. Commun. 15, 5324 (2024).

13. C. Ruis, et al., A systematic nomenclature for mpox viruses causing outbreaks with sustained human-to-human transmission. Nat. Med. 1–5 (2025).

14. F. Kasongo-Mulenda, et al., Clade Ib mpox in the Democratic Republic of the Congo (DRC): Clinical and virological report of the first case in Kinshasa, the capital city. Viruses 17, 1327 (2025).

15. H. Tegally, et al., Dispersal patterns and influence of air travel during the global expansion of SARS-CoV-2 variants of concern. Cell 186, 3277–3290.e16 (2023).

16. Z. Chen, et al., COVID-19 pandemic interventions reshaped the global dispersal of seasonal influenza viruses. Science 386, eadq3003 (2024).

17. M. B. Diamond, et al., Wastewater surveillance of pathogens can inform public health responses. Nat. Med. 28, 1992–1995 (2022).

18. S. V. Scarpino, et al., Socioeconomic bias in influenza surveillance. PLoS Comput. Biol. 16, e1007941 (2020).

19. P. M. Polgreen, et al., Optimizing influenza sentinel surveillance at the state level. Am. J. Epidemiol. 170, 1300–1306 (2009).

20. S. V. Scarpino, N. B. Dimitrov, L. A. Meyers, Optimizing provider recruitment for influenza surveillance networks. PLoS Comput. Biol. 8, e1002472 (2012).

21. S. V. Scarpino, L. A. Meyers, M. A. Johansson, Design strategies for efficient arbovirus surveillance. Emerg. Infect. Dis. 23, 642–644 (2017).

22. P. Nascimento de Lima, et al., The value of environmental surveillance for pandemic response. Sci. Rep. 14, 28935 (2024).

23. M. B. Diamond, E. Yee, M. Bhinge, S. V. Scarpino, Wastewater surveillance facilitates climate change-resilient pathogen monitoring. Sci. Transl. Med. 15, eadi7831 (2023).

24. K.O. Drake, E. Volz, *NBPMscape: Network branching process model for UK MSCAPE* (Github). Manuscript in Preparation.

25. Travel to work by method and distance. Available at: https://www.ons.gov.uk/aboutus/transparencyandgovernance/freedomofinformationfoi/traveltoworkbymethodanddistance [Accessed 11 November 2025].

26. J. Riou, C. L. Althaus, Pattern of early human-to-human transmission of Wuhan 2019 novel coronavirus (2019-nCoV), December 2019 to January 2020. Euro Surveill. 25 (2020).

27. Q. Li, et al., Early transmission dynamics in Wuhan, China, of novel Coronavirus-infected pneumonia. N. Engl. J. Med. 382, 1199–1207 (2020).

28. J. Griffin, et al., Rapid review of available evidence on the serial interval and generation time of COVID-19. BMJ Open 10, e040263 (2020).

29. J. L.-H. Tsui, et al., Genomic assessment of invasion dynamics of SARS-CoV-2 Omicron BA.1. Science 381, 336–343 (2023).

30. D. Brockmann, D. Helbing, The hidden geometry of complex, network-driven contagion phenomena. Science 342, 1337–1342 (2013).

31. The 16th Ebola Virus Disease Outbreak in Bulape Health Zone, Kasai, Democratic Republic of the Congo: A new spillover event from an unknown reservoir host. Virological (2025). Available at: https://virological.org/t/the-16th-ebola-virus-disease-outbreak-in-bulape-health-zone-kasai-democratic-republic-of-the-congo-a-new-spillover-event-from-an-unknown-reservoir-host/1003 [Accessed 6 November 2025].

32. Phylodynamic Analysis. Virological (2020). Available at: https://virological.org/t/phylodynamic-analysis-176-genomes-6-mar-2020/356 [Accessed 6 November 2025].

33. A. N. Cohen, B. Kessel, M. G. Milgroom, Diagnosing SARS-CoV-2 infection: the danger of over-reliance on positive test results. Epidemiology (2020).

34. A. Bivins, R. Morfino, A. Franklin, S. Simpson, W. Ahmed, The lavatory lens: Tracking the global movement of pathogens via aircraft wastewater. Crit. Rev. Environ. Sci. Technol. 54, 321–341 (2024).

35. R. C. Morfino, et al., Notes from the field: Aircraft wastewater surveillance for early detection of SARS-CoV-2 variants - John F. kennedy international airport, New York city, august-September 2022. MMWR Morb. Mortal. Wkly. Rep. 72, 210–211 (2023).

36. J. W. Shingleton, C. J. Lilley, M. J. Wade, Evaluating the theoretical performance of aircraft wastewater monitoring as a tool for SARS-CoV-2 surveillance. *PLOS Glob*. Public Health 3, e0001975 (2023).

37. D. L. Jones, et al., Suitability of aircraft wastewater for pathogen detection and public health surveillance. Sci. Total Environ. 856, 159162 (2023).

38. J. Li, et al., A global aircraft-based wastewater genomic surveillance network for early warning of future pandemics. Lancet Glob. Health 11, e791–e795 (2023).

39. A. J. Kucharski, et al., Effectiveness of isolation, testing, contact tracing, and physical distancing on reducing transmission of SARS-CoV-2 in different settings: a mathematical modelling study. Lancet Infect. Dis. 20, 1151–1160 (2020).

40. K. M. Gostic, A. J. Kucharski, J. O. Lloyd-Smith, Effectiveness of traveller screening for emerging pathogens is shaped by epidemiology and natural history of infection. Elife 4 (2015).

41. Prioritizing diseases for research and development in emergency contexts. Available at: https://www.who.int/activities/prioritizing-diseases-for-research-and-development-in-emergency-contexts [Accessed 10 November 2025].

42. Priority pathogens. Available at: https://cepi.net/priority-pathogens [Accessed 10 November 2025].

43. J. O. Lloyd-Smith, S. J. Schreiber, P. E. Kopp, W. M. Getz, Superspreading and the effect of individual variation on disease emergence. Nature 438, 355–359 (2005).

44. UKHSA Advisory Board: Pathogen Genomics Programme. GOV.UK. Available at: https://www.gov.uk/government/publications/ukhsa-advisory-board-meeting-papers-march-2025/ukhsa-advisory-board-pathogen-genomics-programme [Accessed 22 January 2026].

45. B. X. Tran, et al., Understanding health seeking behaviors to inform COVID-19 surveillance and detection in resource-scarce settings. J. Glob. Health 10, 0203106 (2020).

46. CDC, About. Traveler-based Genomic Surveillance (2025). Available at: https://www.cdc.gov/traveler-genomic-surveillance/about/index.html [Accessed 10 November 2025].

47. pgfgleam: Probability generating function methodology for epidemic metapopulation models (Github).

48. D. Balcan, et al., Modeling the spatial spread of infectious diseases: the GLobal Epidemic and Mobility computational model. J. Comput. Sci. 1, 132–145 (2010).

49. M. Chinazzi, et al., A multiscale modeling framework for Scenario Modeling: Characterizing the heterogeneity of the COVID-19 epidemic in the US. Epidemics 47, 100757 (2024).

50. J. T. Davis, et al., Cryptic transmission of SARS-CoV-2 and the first COVID-19 wave. Nature 600, 127–132 (2021).

51. G. Zagatti, et al., Extending JumpProcesses.jl for fast point process simulation with time-varying intensities. JuliaCon Proceedings 6, 133 (2024).

52. Data and analysis. Available at: https://www.caa.co.uk/data-and-analysis/ [Accessed 26 November 2025].

53. Origin-destination data, England and Wales: Census 2021 - Nomis - Official Census and Labour Market Statistics. Available at: https://www.nomisweb.co.uk/sources/census_2021_od [Accessed 26 November 2025].

54. J. Mossong, et al., Social contacts and mixing patterns relevant to the spread of infectious diseases. PLoS Med. 5, e74 (2008).

55. L. Ferretti, et al., Quantifying SARS-CoV-2 transmission suggests epidemic control with digital contact tracing. Science 368, eabb6936 (2020).

56. Report of the WHO-China Joint Mission on coronavirus disease 2019 (COVID-19) (2020).

57. E. S. Knock, et al., Key epidemiological drivers and impact of interventions in the 2020 SARS-CoV-2 epidemic in England. Sci. Transl. Med. 13, eabg4262 (2021).

58. T. Charalampous, et al., Routine metagenomics service for ICU patients with respiratory infection. Am. J. Respir. Crit. Care Med. 209, 164–174 (2024).

59. W. Gu, et al., Rapid pathogen detection by metagenomic next-generation sequencing of infected body fluids. Nat. Med. 27, 115–124 (2021).

60. N. C. Stenseth, et al., How to avoid a local epidemic becoming a global pandemic. Proc. Natl. Acad. Sci. U. S. A. 120, e2220080120 (2023).

