## Supplementary for "A multi-scale model to evaluate airport wastewater surveillance and ICU genomic monitoring for pandemic preparedness"

#### Supplementary Materials

##### Relationship between air travel, detection timing, and local case burden

In our SARS-CoV-2-like example, we fix $R_0$=2 for each country; therefore the only differential as we vary outbreak origins is from global dissemination across the airline network. Moreso, in early scenarios, given the delay of secondary countries becoming large exporters of importations to EW (the delay for other countries to get the outbreak, and then latent period lags), we can approximate the exponential growth of importations to be β; such that $\lambda(t)$ = E[Daily Importations into EW](t) $\approx mI_0\, e^{\kappa t}$, where m is the travel rate (assumed to be constant)

Let $t_A$ and $t_D$ denote the time of arrival (the time at which the number of cumulative importations exceeds 1), and the AWW detection time, respectively.

Then we can express the difference in these times as

$$|t_D - t_A| \approx \frac{1}{\beta} \log(K_D(s) / mI_0),$$

where $K_D(s)$ is the threshold of importations required to reach detection given an outbreak origin of country s. Since the probability of detection is fixed to 0.16 irrespective of outbreak origin in our current approach, we expect $K_D(s)$ to be similar across outbreak origins, and so the above equation is approximately constant. This is highlighted in Fig. 2A**,** where despite a large variance in arrival and WW detection times, their difference varies by only a few days across all scenarios.

##### Mathematical Derivation of Importation Distributions Linking Global and Local Models

We can model the cumulative importations as above, such that C(t) then follows a Poisson distribution.

$$C(t) \sim \mathrm{Poisson}\left(\int_0^t \lambda(s)ds\right) = \mathrm{Poisson}\left(\frac{1}{\kappa} mI_0[e^{\kappa t} - 1]\right).$$

As $\lambda(t)$ is approximately deterministic early on, the variance is equal to the mean (i.e. Var[C(t)] = E[C(t)], Supplementary Fig. 2A-B) which is why we use a Poisson model in the early stages of an outbreak. As the epidemic spreads and global dissemination occurs, EW receives importations from a much larger number of countries; and so, overdispersion is present (Supplementary Fig. 2C). The total rate, $\Lambda(t) = \Sigma\, m_{i,EW} I_i(t)$ becomes a random variable, as each country's infection level $I_i(t)$ depends on its own stochastic seeding history.

Following the law of total variance, $Var[Y_{EW}] = E[\Lambda] + Var[\Lambda]$

Var[Λ] represents the uncertainty in the importation rate across different epidemic realisations. In particular, when Λ is Gamma-distributed, the marginal distribution of importations $Y_{EW}$ follows a Negative Binomial (NegBin) distribution:

$$Y_{EW} \sim \text{NegBin}(r = \alpha, p = \frac{\beta}{1 + \beta})$$

For the results throughout the manuscript, rather than fitting a NegBin distribution to the cumulative results and sampling from the resulting posterior, we take Poisson samples of the instantaneous mean importation rate $\Lambda(t)$ at each time step. This approach is computationally efficient as it allows us to rely solely on the cumulants extraction of the global model, which utilises FFT methods for quick computation. While this assumes the variance is locally Poisson (Var = Mean), it captures the primary source of early-outbreak stochasticity. We note that, for later epidemic stages, this can lose the variance in individual samples, since the Poisson sampling loses the overdispersion (Var[$\Lambda$]) present in the NegBin distribution. However, since this study is designed for early epidemic scenarios, we do not think that this is a major issue.

### Supplementary Figures and Tables

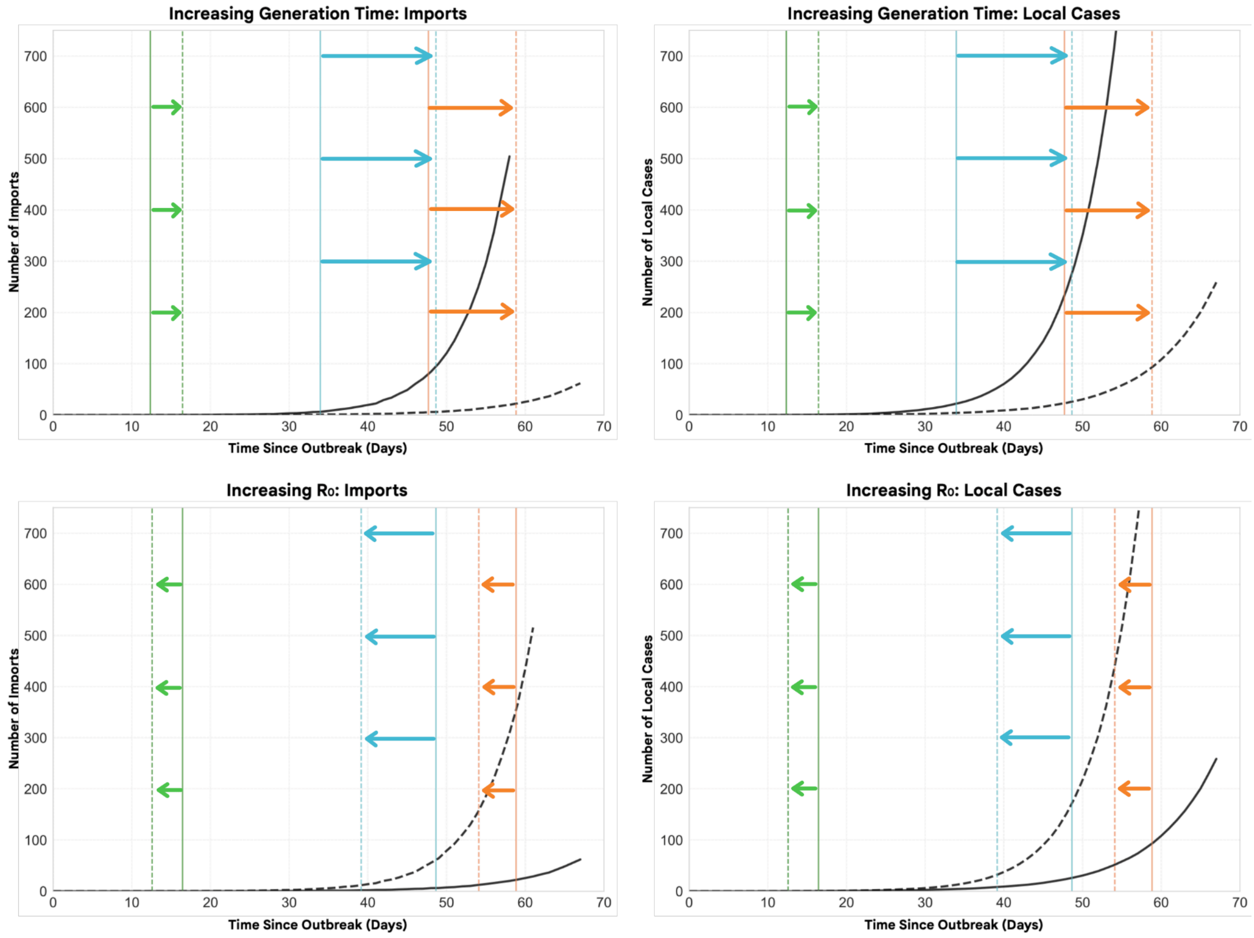


***Supplementary Figure 1:*** *Plots showing the effects of changing epidemiological parameters (basic reproduction number, generation time) on the mean importation array, and the mean EW local cases array. Vertical lines correspond to arrival time (green), AWW detection (blue) and ICU detection time (ICU), with solid lines representing the initial parameters, and the dashed lines representing the increased parameter; as specified in the panel title. We arbitrarily plot for an outbreak origin of the United Arab Emirates, with initial parameters of (R0, generation time) = (2.0, 4.0). An increase in the basic reproduction number corresponds to an increase from 2.0 to 2.5, and an increase in generation time corresponds to an increase from 4.0 to 6.0.*

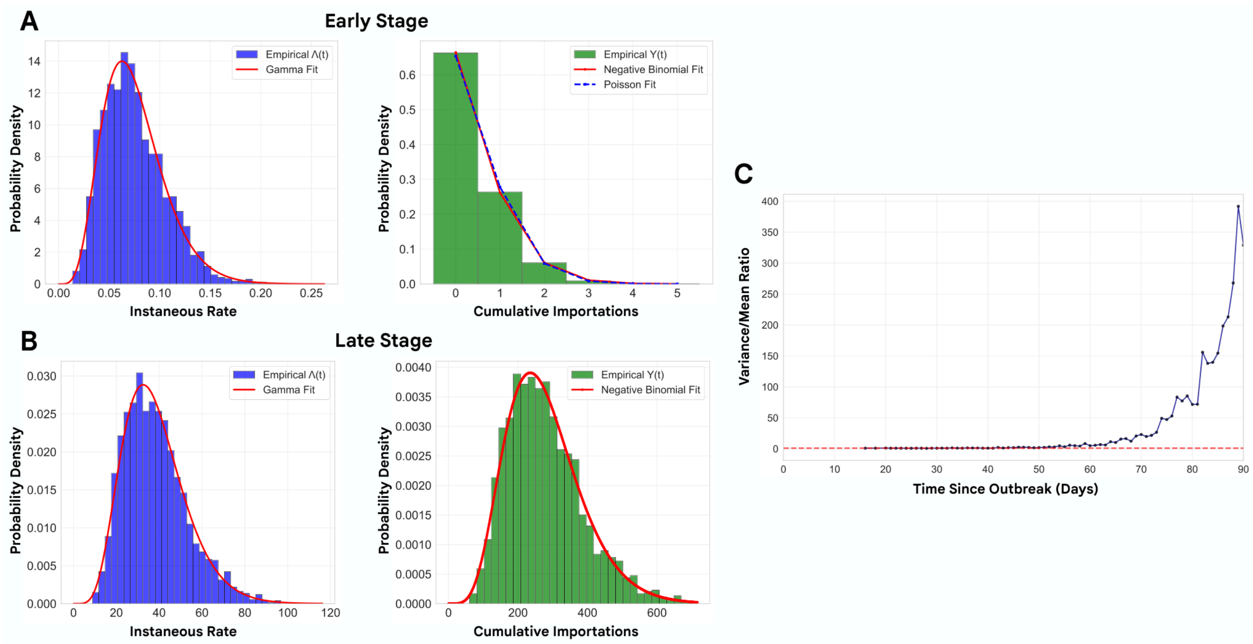


***Supplementary Figure 2****. Monte Carlo simulation of importations through a synthetic 6-country network (n=1000 realisations).* ***(A)*** *Early phase (t=30d): Instantaneous rate Λ(t) follows Gamma distribution, while cumulative importations Y(t) fitted to NegBin distribution effectively collapses to a Poisson distribution (Var/Mean=1.11); justifying simplified Poisson seeding in initial outbreak stages.* ***(B)*** *Late phase (t=75d): Λ(t) remains Gamma-distributed but with larger variance. As secondary transmission pathways within the network become viable, the variance in the importation rate increases. Cumulative importations Y(t) show strong overdispersion (Var/Mean=41.6), where the Negative Binomial fit becomes essential to capture the increased frequency of high-volume importation events. Red curves show fitted distributions* ***(C)*** *Sampling validation: Variance-to-mean ratio over time demonstrates the natural emergence of overdispersion as infections spread across the network. The convergence to 1 (red line) for early time highlights the Poisson seeding viability.*

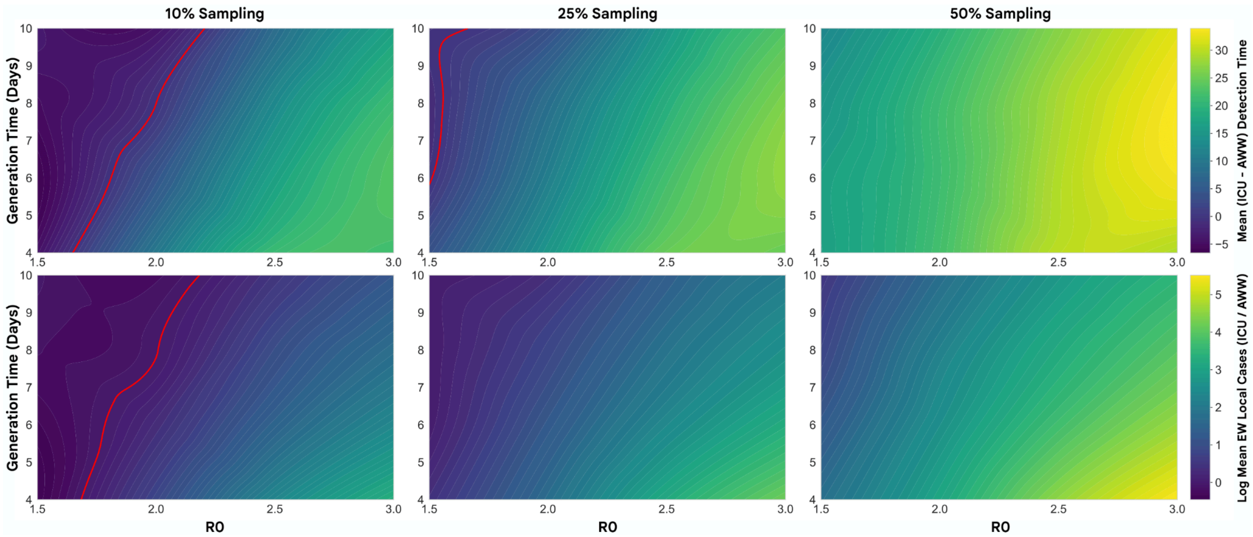


***Supplementary Figure 3:*** *Heatmaps showing the estimated difference in mean detection time (top row) and difference in mean EW local cases at time of detection (bottom row) across parameter space; varying $R_0$ (x-axis), mean generation time (y-axis), and aircraft sampling rate (10%, 25%, 50% across columns), and ICU sampling rate of 10%. Similar to Figure* ***3B)****, but now assuming an individual-level AWW detection probability of 8%. The red zero contour line on the heatmaps represents when the AWW and ICU are equal (in mean detection times and local cases at time of detection); points to the left of the red contour line correspond to when ICU outperforms AWW surveillance.*

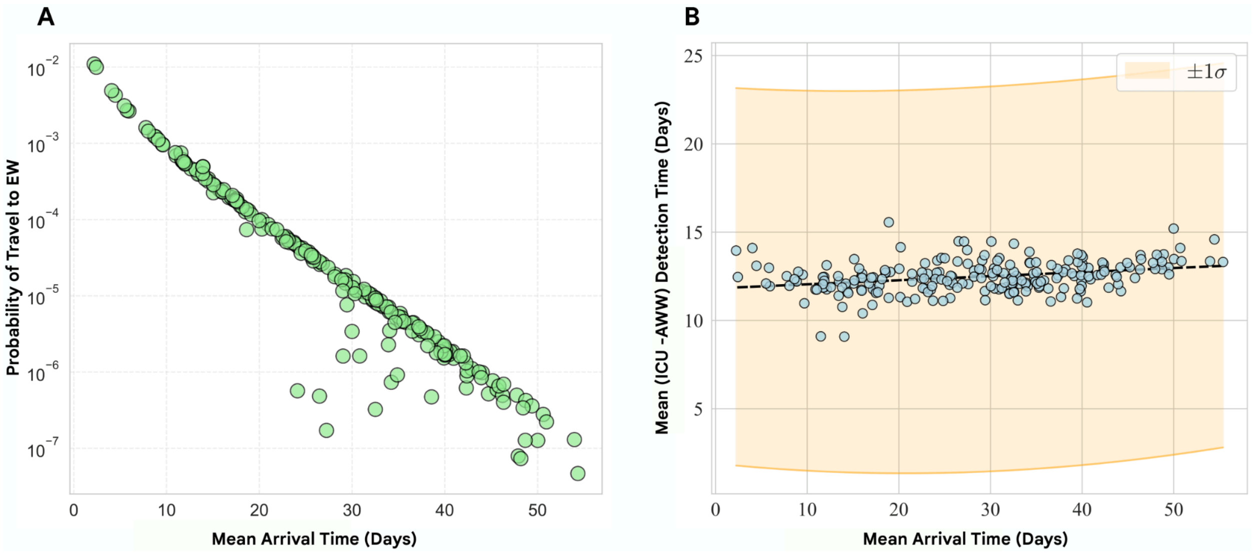


***Supplementary Figure 4: (A)*** *Probability of travelling to EW as a function of arrival time for outbreak origins in different countries. Each scatter point represents a country, with a clear negative log relationship between the two factors. Points that empirically lie below the regression line tend to be countries that have weak or no direct links to EW, but strong connections to other countries which have*

*strong travel themselves to EW; resulting in dominance of secondary importers.* ***(B)*** *Distribution of mean detection time differences (ICU - AWW) across 100 simulations of the local model for each country outbreak origin. Points represent simulation means, for a given origin country, with ±1σ error ribbons representing sample variation.*

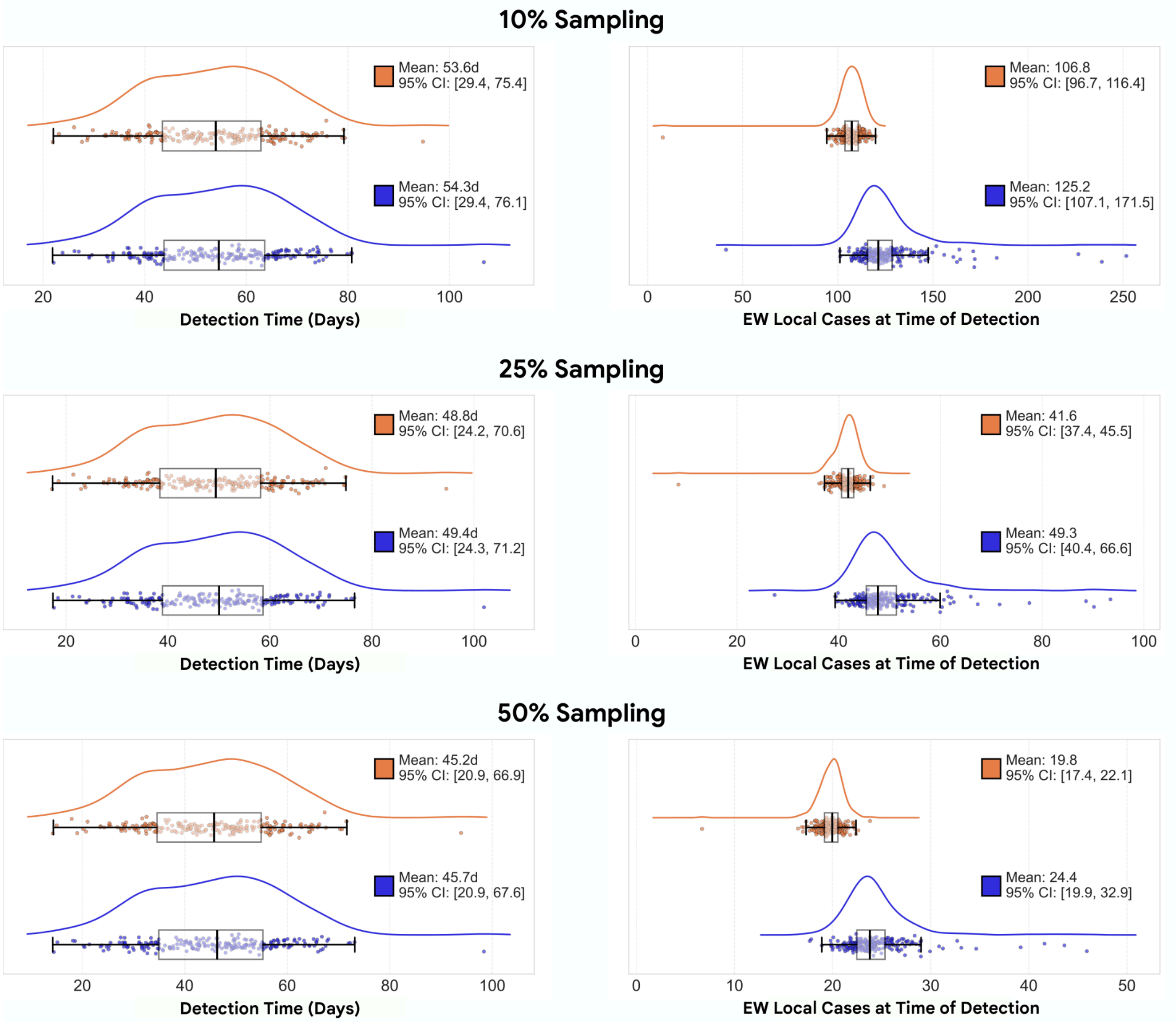


***Supplementary Figure 5:*** *Comparison of AWW mean detection times when we test flights from all origins (orange), and testing flights from only the top 3 origins (dark blue), for a SARS-CoV-2 like virus. Each row corresponds to 10, 25 and 50% aircraft sampling (for T3 this means sampling x% of the incoming aircrafts from the top 3 countries which contribute to import flux, given each country as a candidate outbreak origin). The left column compares the detection time between testing all flights and only a select few across all outbreak origins (each individual scatter corresponding to the mean of 100 samples given a country as OL) with box plots indicating median, interquartile range, and 95% confidence intervals displayed. The right column then plots the number of local cases at detection time. These results highlight that the majority of the early importation flux is derived from a small number of origins.*

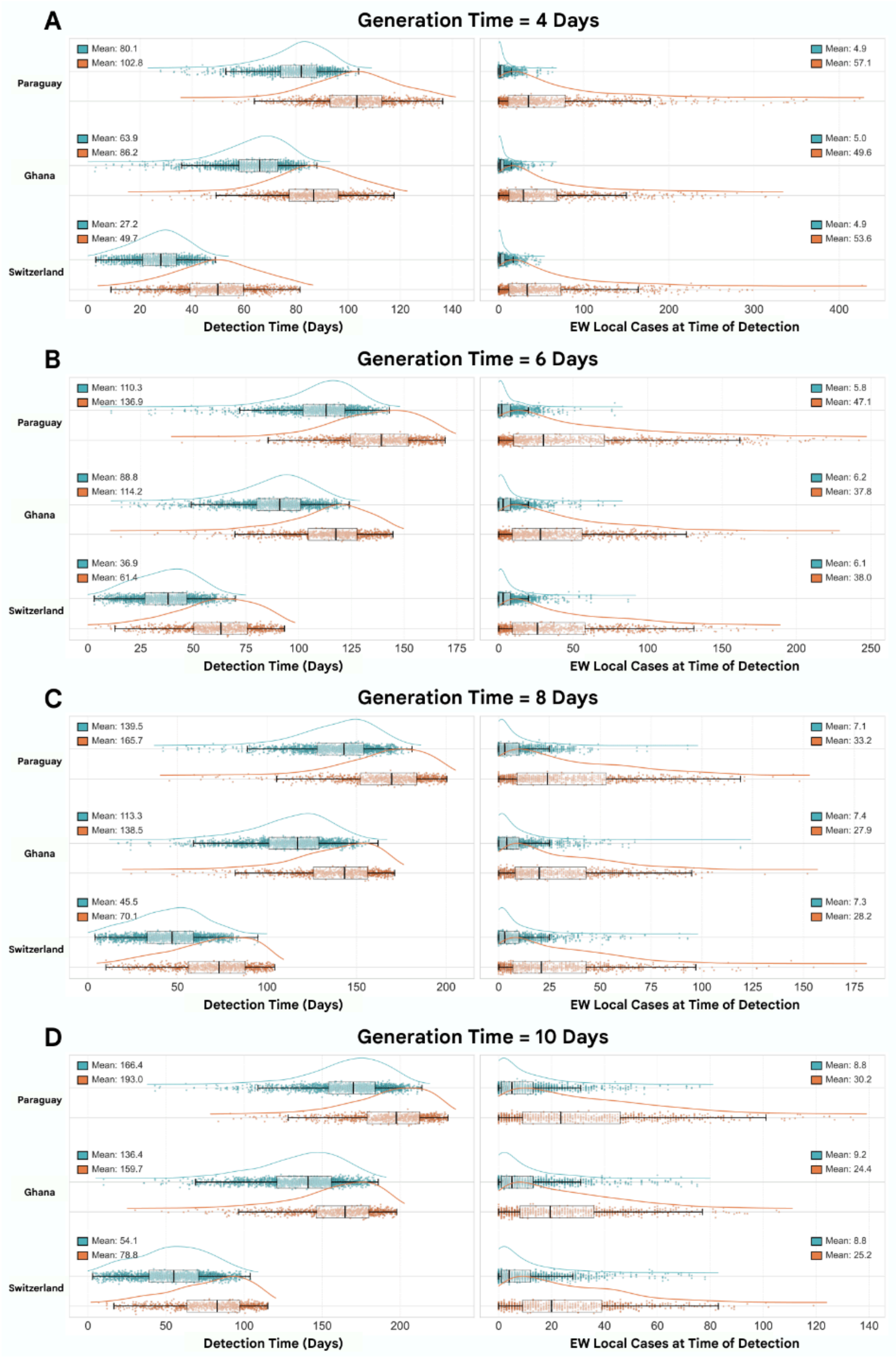


***Supplementary Figure 6:*** *Global scenario - $R_0$ = 1.5, 50% Aircraft Sampling Rate.* ***(A)*** *Generation Time = 4 days.* ***(B)*** *Generation Time = 6 days.* ***(C)*** *Generation Time = 8 days.* ***(D)*** *Generation Time = 10 days.*

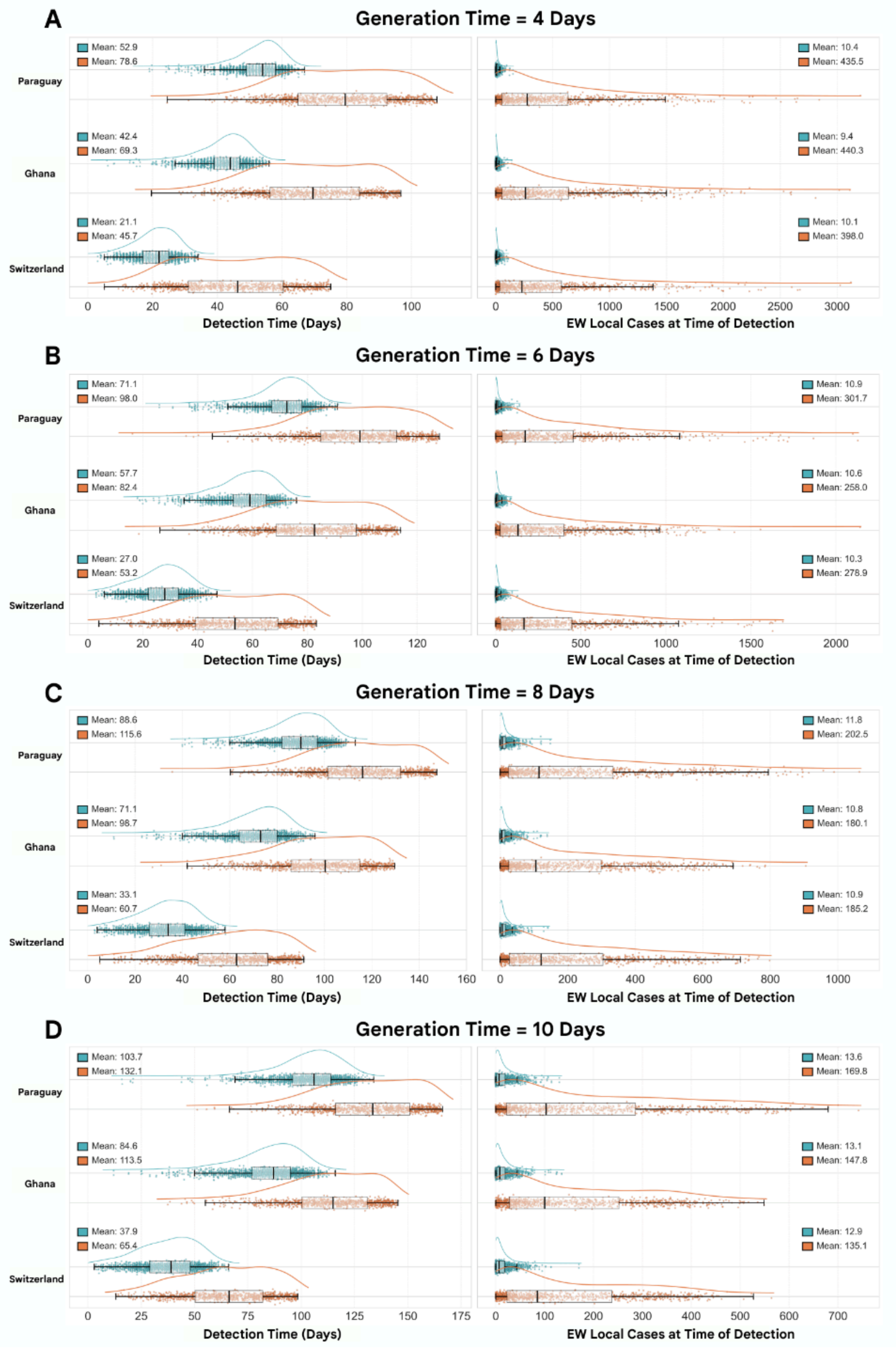


***Supplementary Figure 7:*** *Global scenario - $R_0$ = 2.0, 50% Aircraft Sampling Rate.* ***(A)*** *Generation Time = 4 days.* ***(B)*** *Generation Time = 6 days.* ***(C)*** *Generation Time = 8 days.* ***(D)*** *Generation Time = 10 days.*

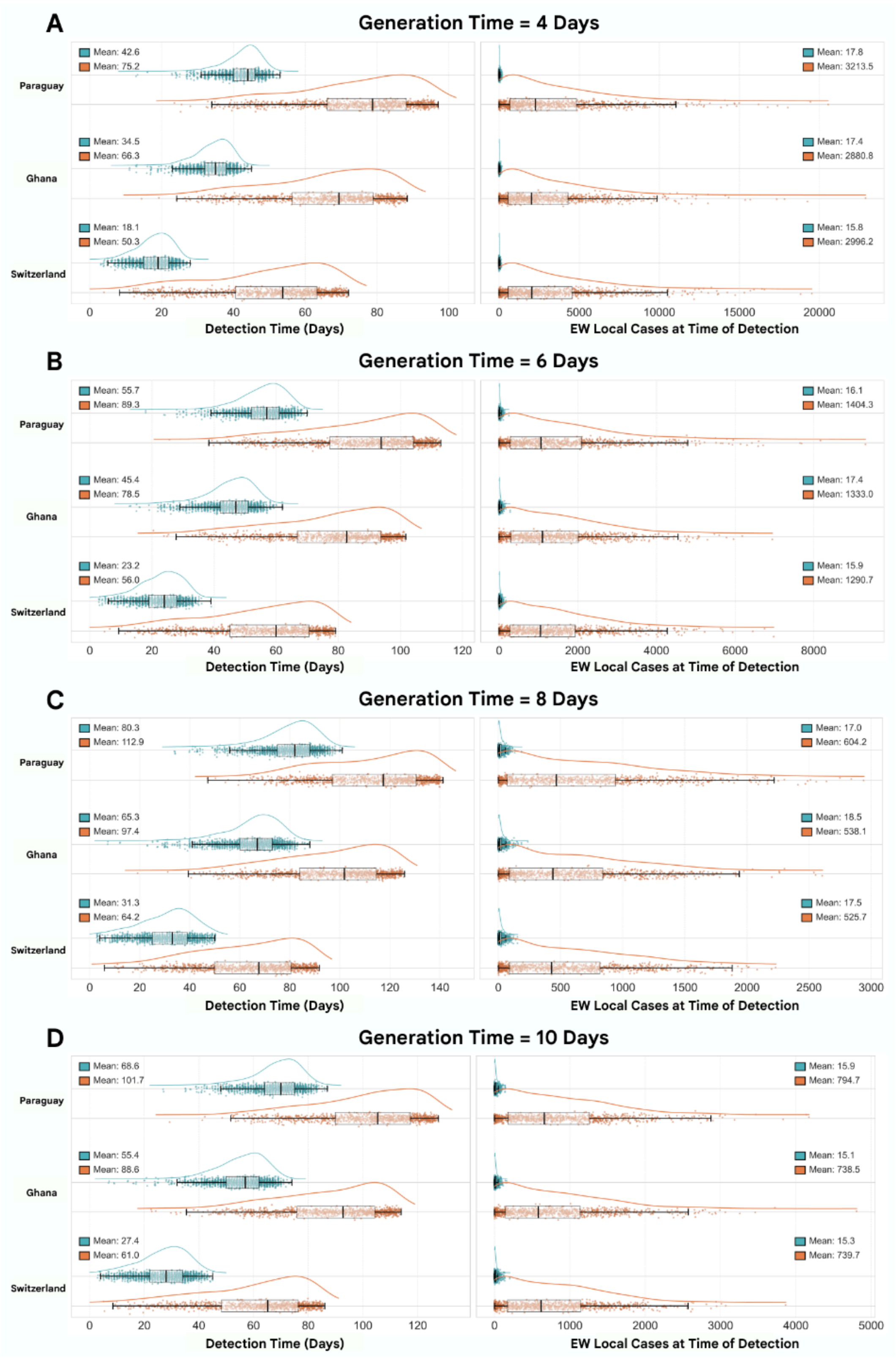


***Supplementary Figure 8:*** *Global scenario - $R_0$ = 2.5, 50% Aircraft Sampling Rate.* ***(A)*** *Generation Time = 4 days.* ***(B)*** *Generation Time = 6 days.* ***(C)*** *Generation Time = 8 days.* ***(D)*** *Generation Time = 10 days.*

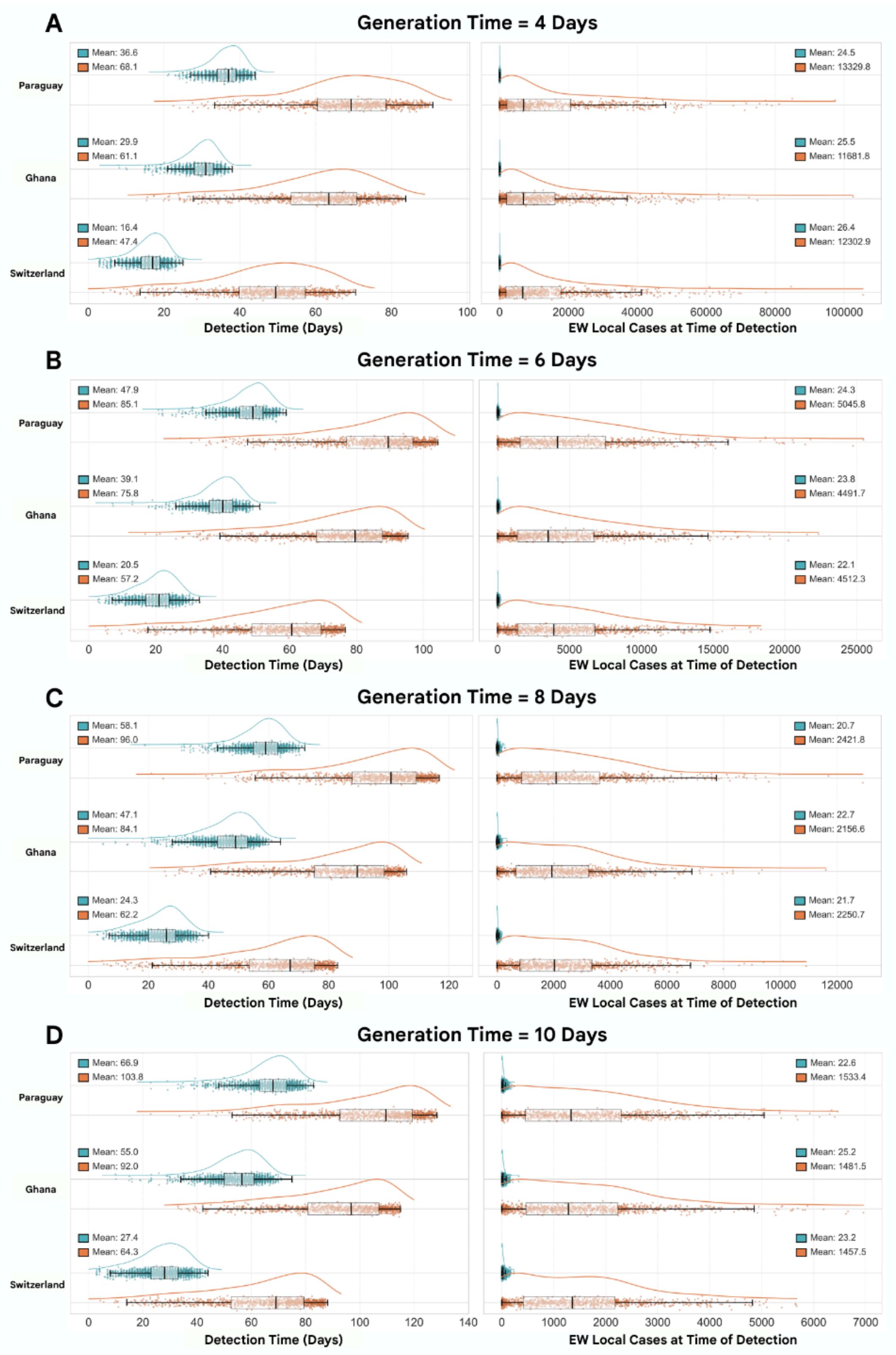


***Supplementary Figure 9:*** *Global scenario - $R_0$ = 3.0, 50% Aircraft Sampling Rate.* ***(A)*** *Generation Time = 4 days.* ***(B)*** *Generation Time = 6 days.* ***(C)*** *Generation Time = 8 days.* ***(D)*** *Generation Time = 10 days.*

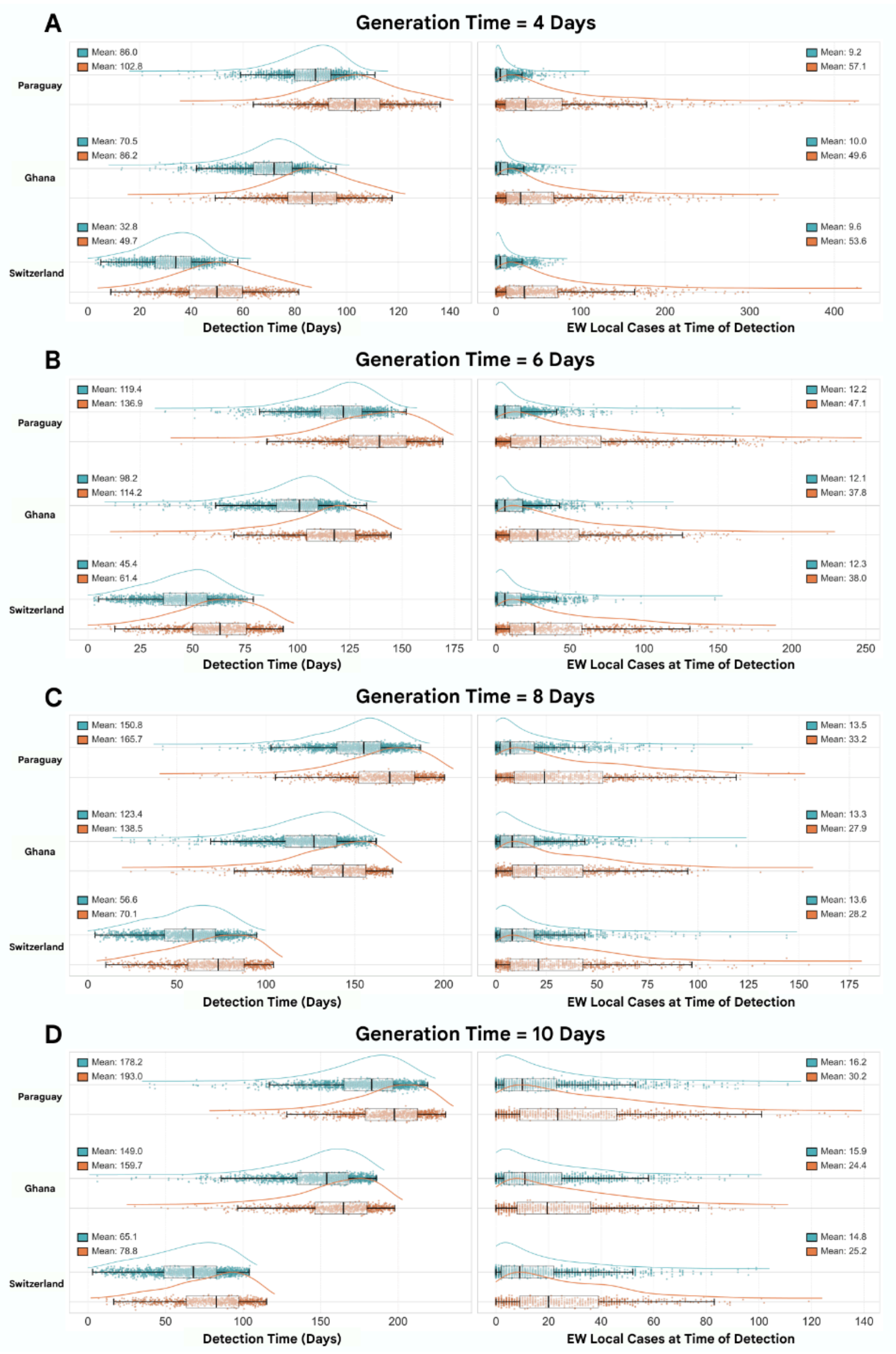


***Supplementary Figure 10:*** *Global scenario - $R_0$ = 1.5, 25% Aircraft Sampling Rate.* ***(A)*** *Generation Time = 4 days.* ***(B)*** *Generation Time = 6 days.* ***(C)*** *Generation Time = 8 days.* ***(D)*** *Generation Time = 10 days.*

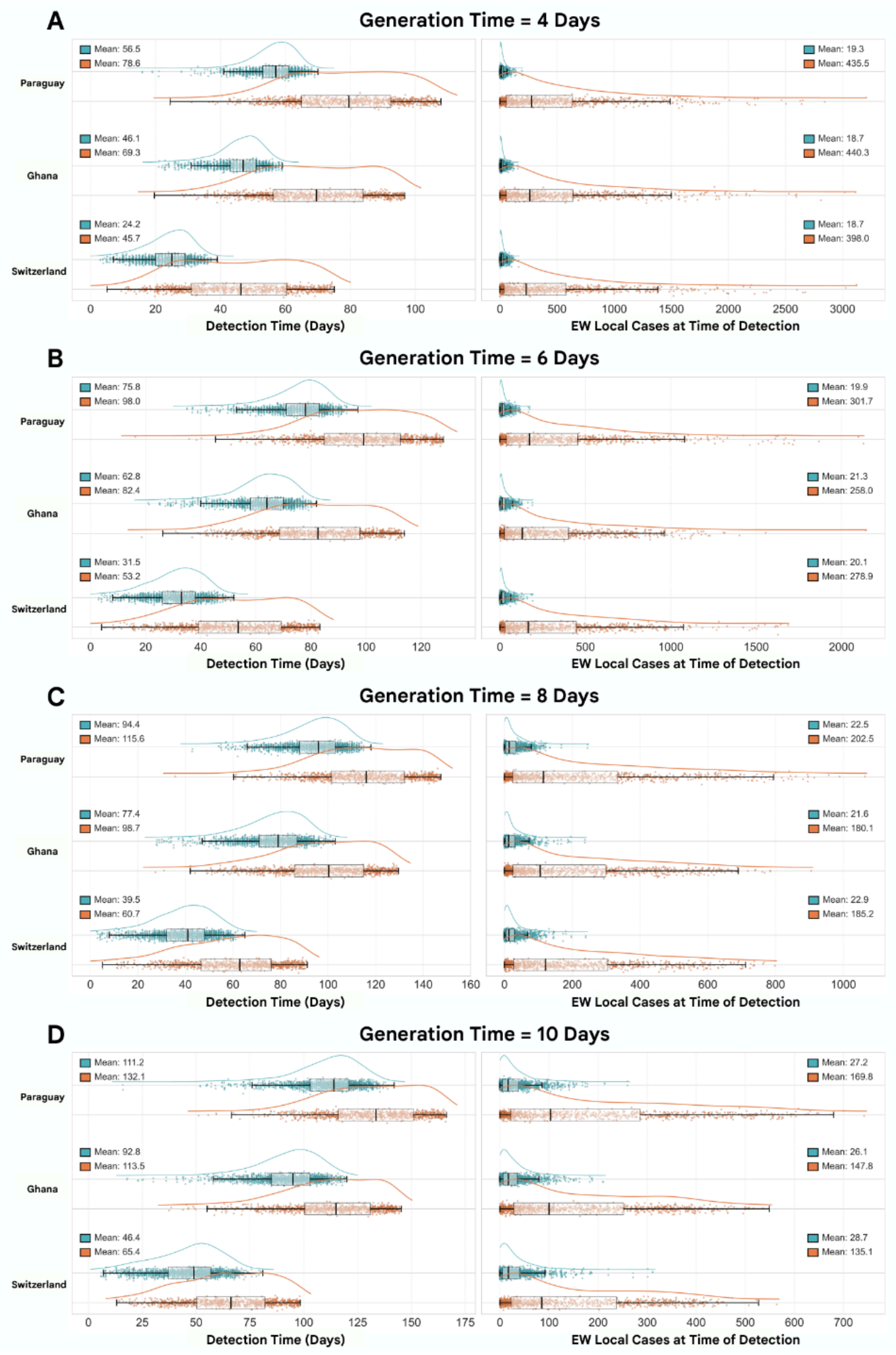


***Supplementary Figure 11:*** *Global scenario - $R_0$ = 2.0, 25% Aircraft Sampling Rate.* ***(A)*** *Generation Time = 4 days.* ***(B)*** *Generation Time = 6 days.* ***(C)*** *Generation Time = 8 days.* ***(D)*** *Generation Time = 10 days.*

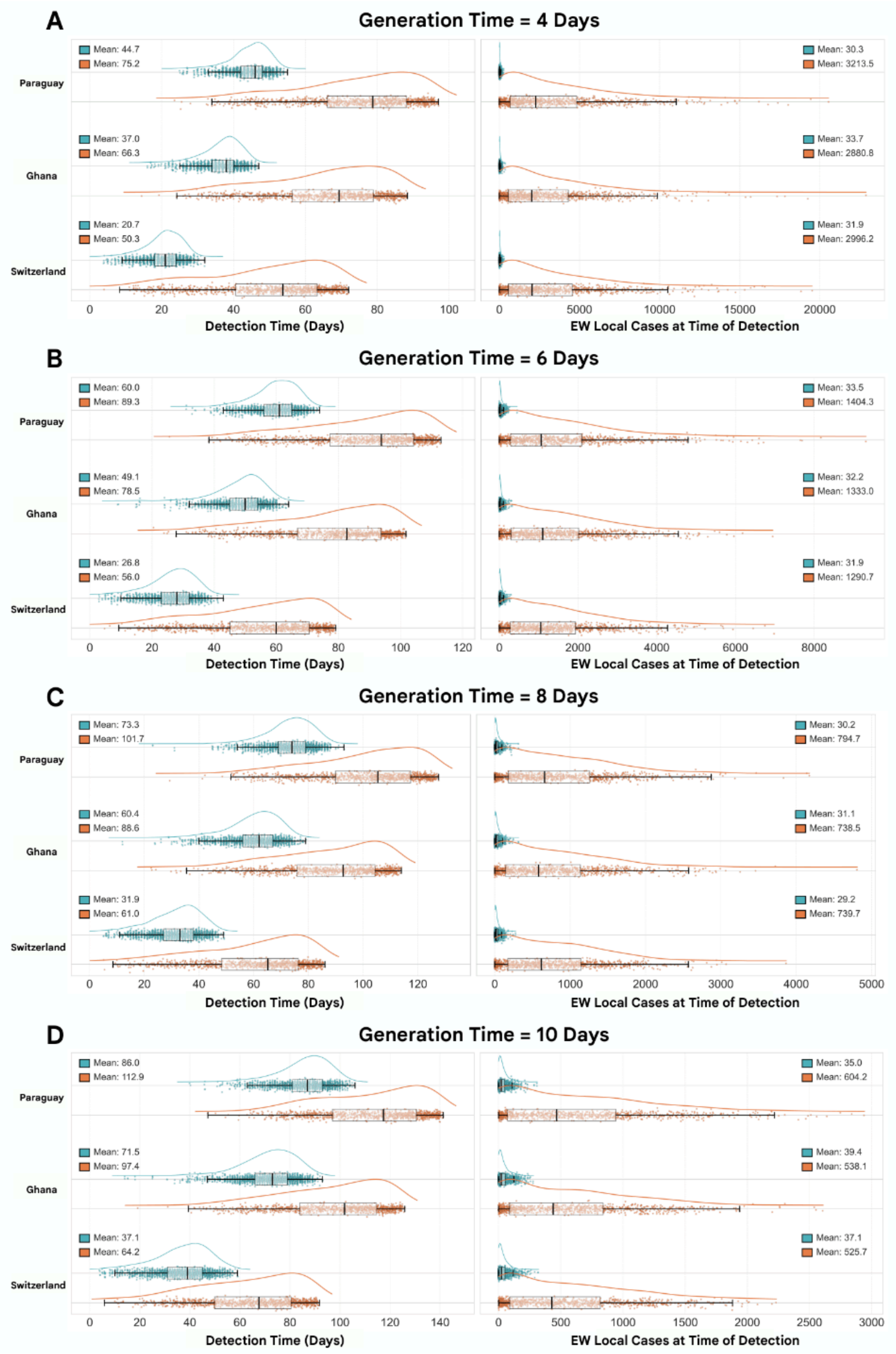


***Supplementary Figure 12:*** *Global scenario - $R_0$ = 2.5, 25% Aircraft Sampling Rate.* ***(A)*** *Generation Time = 4 days.* ***(B)*** *Generation Time = 6 days.* ***(C)*** *Generation Time = 8 days.* ***(D)*** *Generation Time = 10 days.*

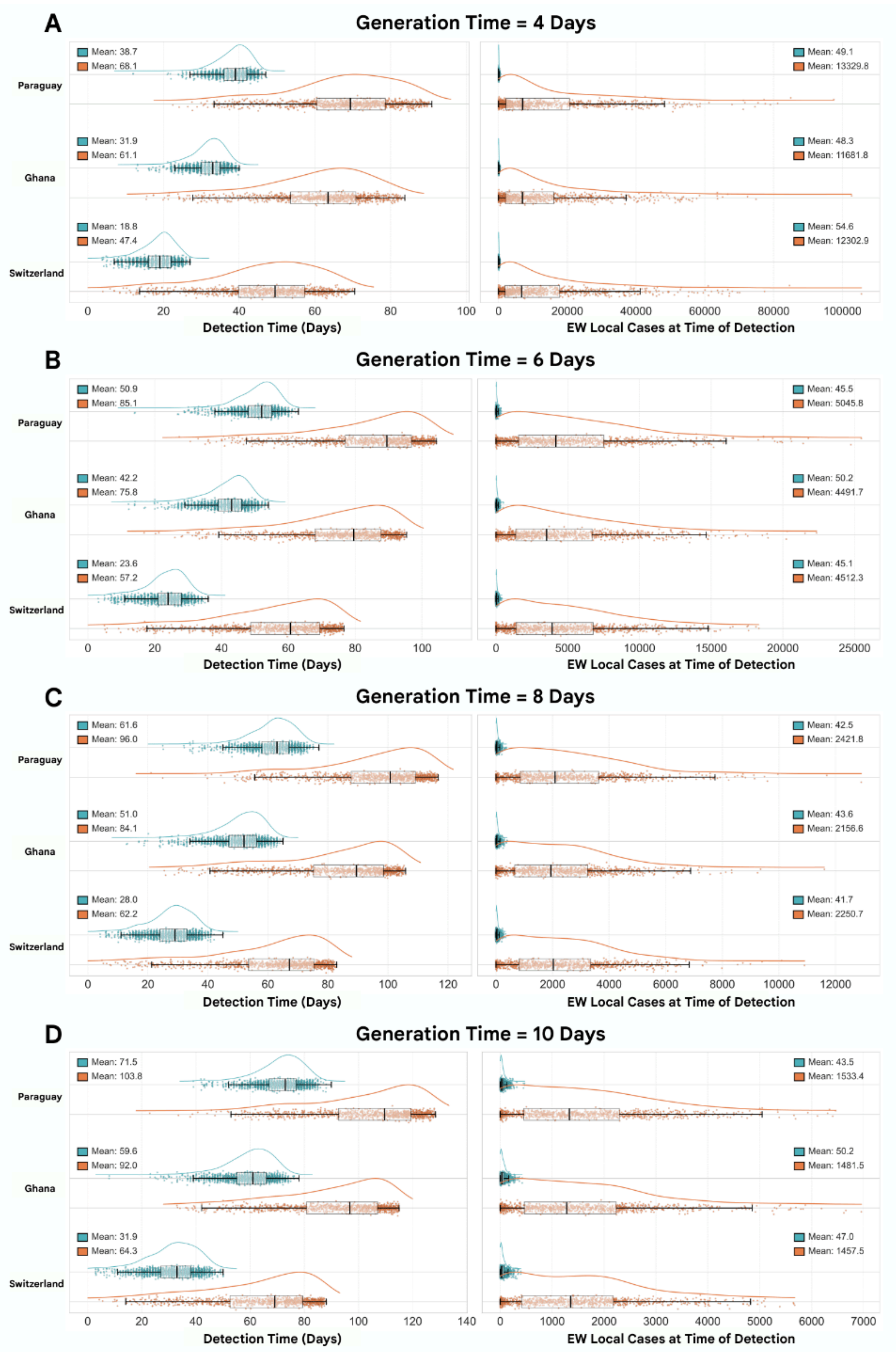


***Supplementary Figure 13:*** *Global scenario - $R_0$ = 3.0, 25% Aircraft Sampling Rate.* ***(A)*** *Generation Time = 4 days.* ***(B)*** *Generation Time = 6 days.* ***(C)*** *Generation Time = 8 days.* ***(D)*** *Generation Time = 10 days.*

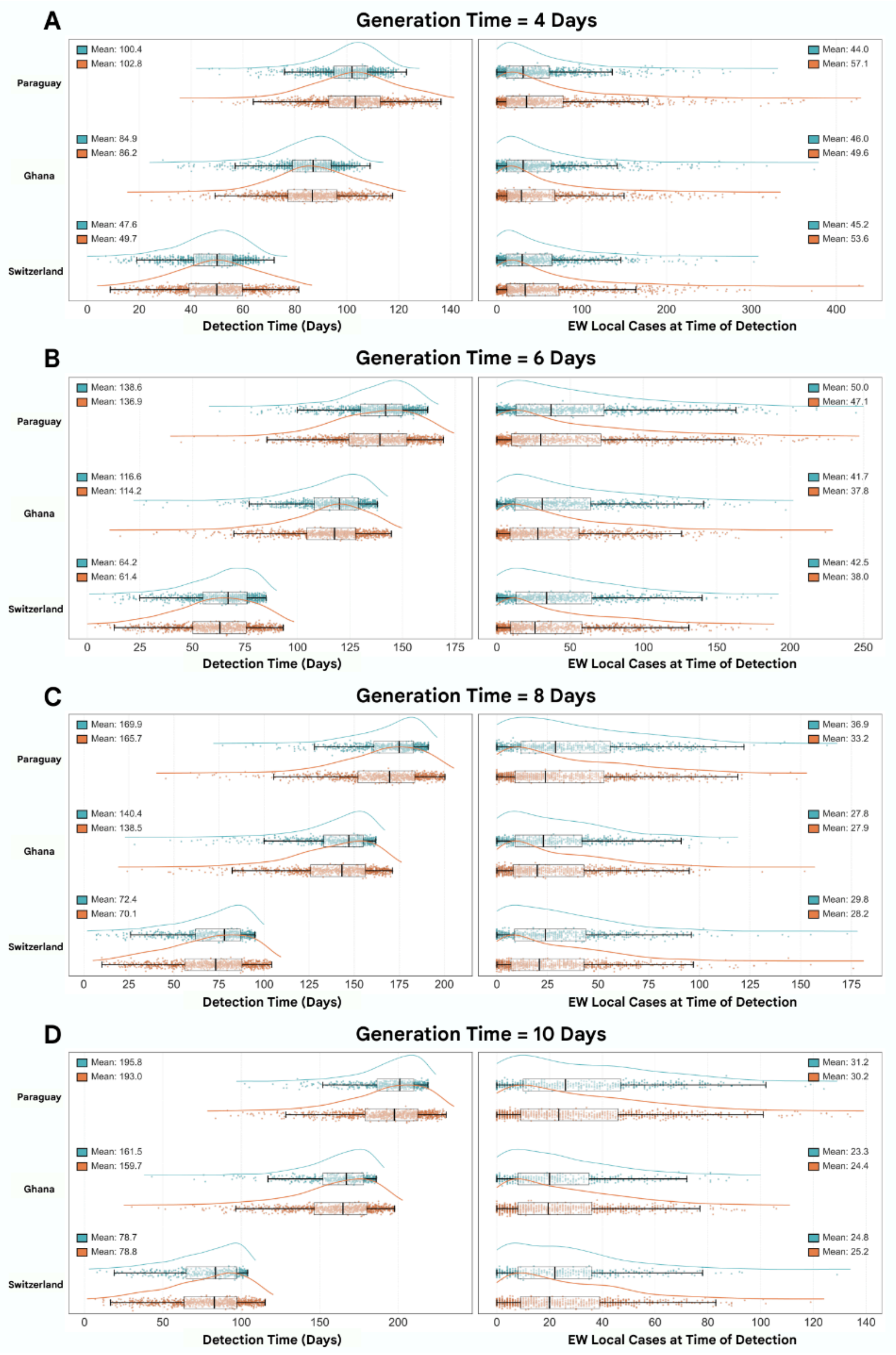


***Supplementary Figure 14:*** *Global scenario - $R_0$ = 1.5, 10% Aircraft Sampling Rate.* ***(A)*** *Generation Time = 4 days.* ***(B)*** *Generation Time = 6 days.* ***(C)*** *Generation Time = 8 days.* ***(D)*** *Generation Time = 10 days.*

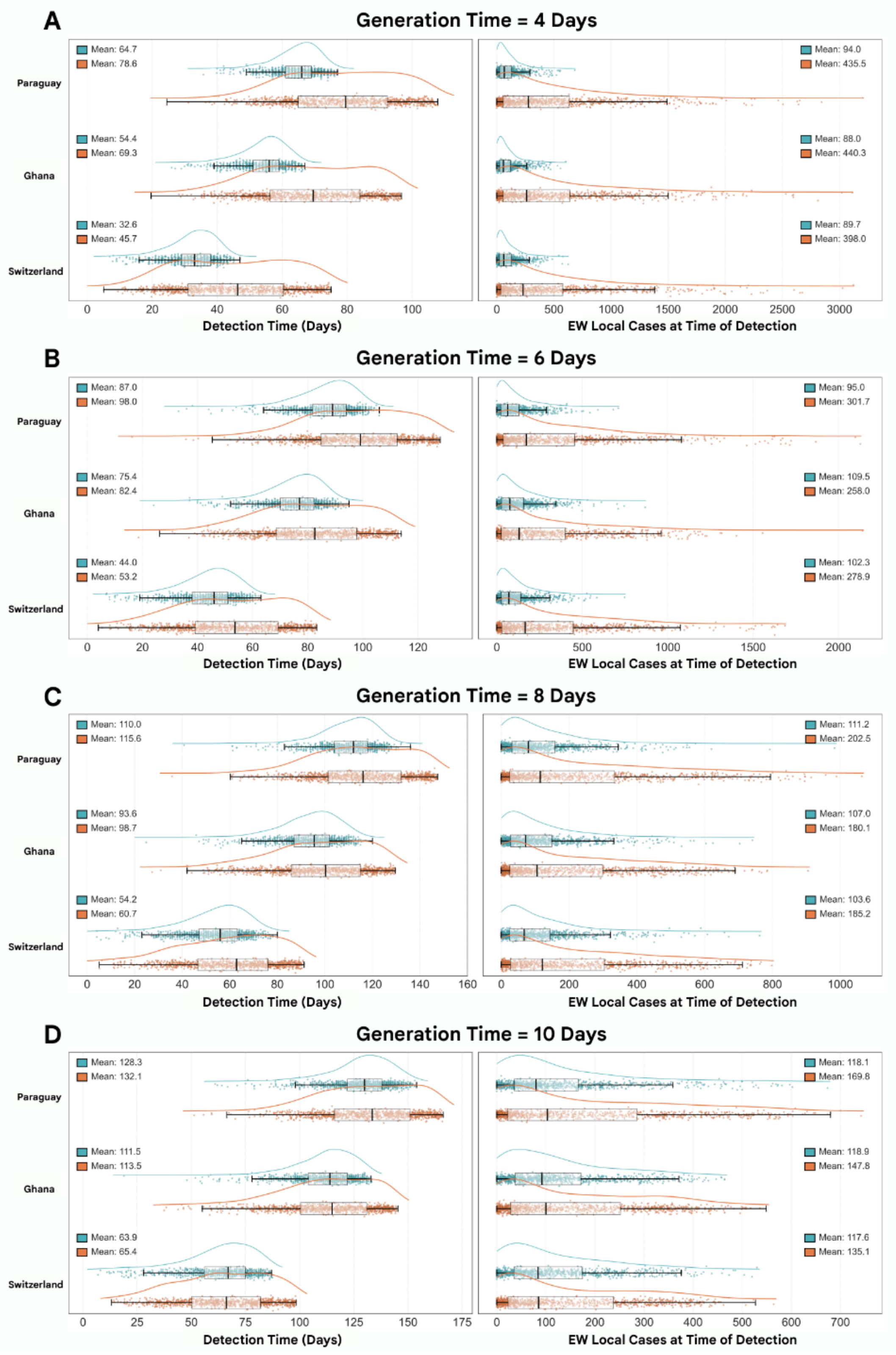


***Supplementary Figure 15:*** *Global scenario - $R_0$ = 2.0, 10% Aircraft Sampling Rate.* ***(A)*** *Generation Time = 4 days.* ***(B)*** *Generation Time = 6 days.* ***(C)*** *Generation Time = 8 days.* ***(D)*** *Generation Time = 10 days.*

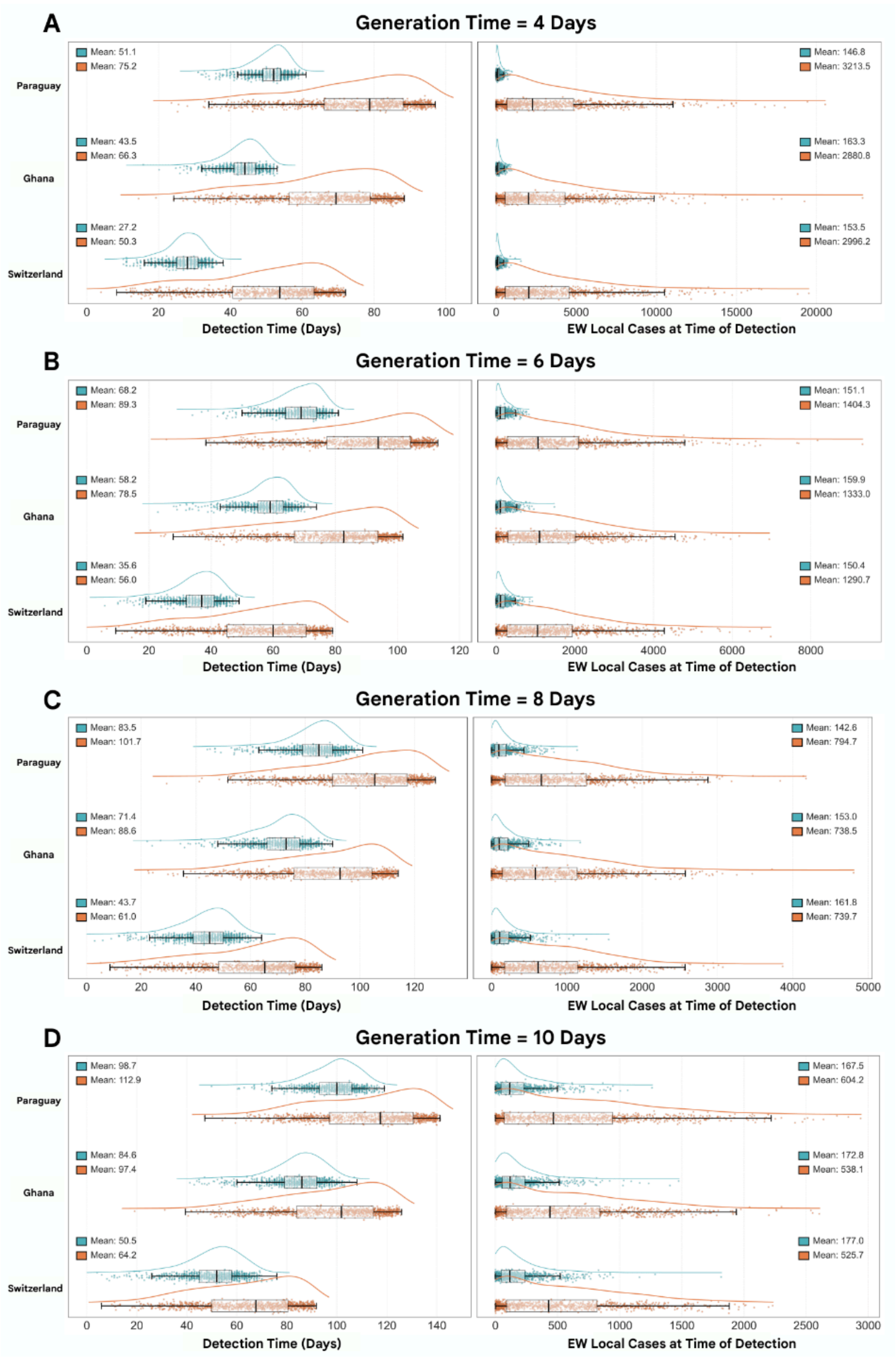


***Supplementary Figure 16:*** *Global scenario - $R_0$ = 2.5, 10% Aircraft Sampling Rate.* ***(A)*** *Generation Time = 4 days.* ***(B)*** *Generation Time = 6 days.* ***(C)*** *Generation Time = 8 days.* ***(D)*** *Generation Time = 10 days.*

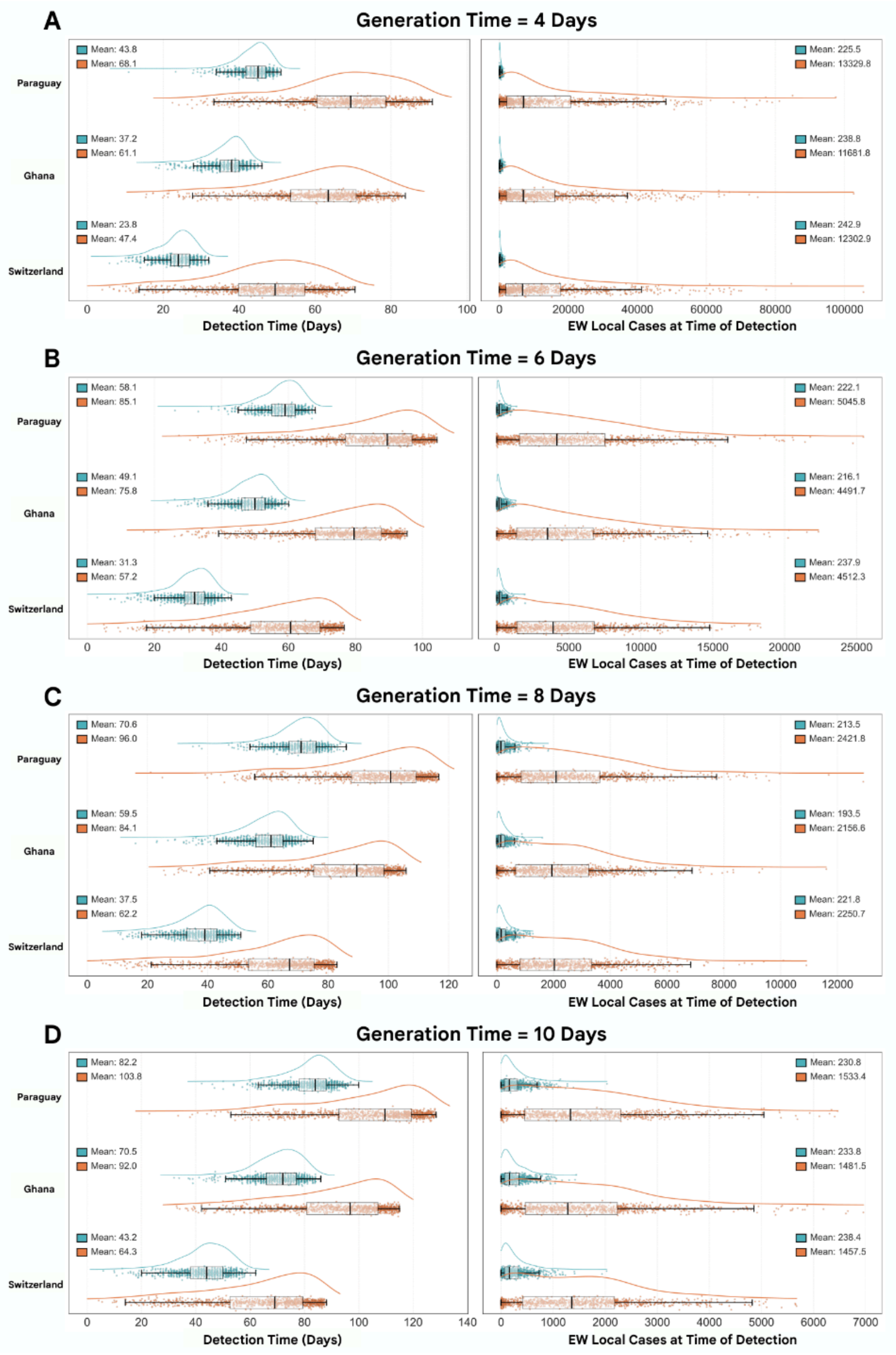


***Supplementary Figure 17:*** *Global scenario - $R_0$ = 3.0, 10% Aircraft Sampling Rate.* ***(A)*** *Generation Time = 4 days.* ***(B)*** *Generation Time = 6 days.* ***(C)*** *Generation Time = 8 days.* ***(D)*** *Generation Time = 10 days.*

| Method (Aircraft Sampling %) | $p_{det}$ | Mean Detection Time (Days) | | EW Local Cases at Time of Detection | |
|---|---|---|---|---|---|
| | | Means (95% CI) | Bootstrap (95% PI) | Means (95% CI) | Bootstrap (95% PI) |
| **ICU** | | 84.3: (81.4, 87.2) | 84.3: (31.7, 134.2) | 55.7: (54.7, 56.7) | 55.8: (0.0, 241.0) |
| **AWW (10%)** | **8%** | 88.7: (85.9, 91.4) | 88.7: (41.0, 132.0) | 87.9: (86.5, 89.3) | 88.1: (0.0, 328.0) |
| **AWW (25%)** | **8%** | 80.4: (77.6, 83.2) | 80.4: (32.0, 124.0) | 36.5: (35.9, 37.1) | 36.4: (0.0, 141.0) |
| **AWW (50%)** | **8%** | 67.9: (65.0, 70.7) | 67.8: (19.0, 111.0) | 9.4: (9.3, 9.6) | 9.4: (0.0, 44.0) |
| **AWW (100%)** | **8%** | 61.9: (59.0, 64.7) | 61.7: (15.0, 106.0) | 4.8: (4.7, 4.9) | 4.8: (0.0, 26.0) |
| **AWW (10%)** | **16%** | 82.3: (79.5, 85.1) | 82.3: (34.0, 126.0) | 45.5: (44.8, 46.3) | 45.9: (0.0, 176.0) |
| **AWW (25%)** | **16%** | 67.9: (65.0, 70.7) | 67.7: (19.0, 111.0) | 9.4: (9.3, 9.6) | 9.4: (0.0, 45.0) |
| **AWW (50%)** | **16%** | 61.9: (59.0, 64.7) | 62.0: (15.0, 106.0) | 4.8: (4.7, 4.9) | 4.8: (0.0, 26.0) |
| **AWW (100%)** | **16%** | 61.6: (58.7, 64.4) | 61.4: (15.0, 105.0) | 4.7: (4.6, 4.8) | 4.7: (0.0, 25.0) |

***Supplementary Table 1****: Global scenario - $R_0$ = 1.5, Generation Time = 4.0 days.*

| Method (Aircraft Sampling %) | $p_{det}$ | Mean Detection Time (Days) | | EW Local Cases at Time of Detection | |
|---|---|---|---|---|---|
| | | Means (95% CI) | Bootstrap (95% PI) | Means (95% CI) | Bootstrap (95% PI) |
| **ICU** | | 110.1: (106.2, 114.0) | 110.3: (40.3, 174.1) | 41.6: (40.9, 42.3) | 41.8: (0.0, 150.0) |
| **AWW (10%)** | **8%** | 116.6: (112.7, 120.6) | 116.6: (50.0, 177.0) | 58.2: (57.0, 59.4) | 58.4: (0.0, 165.0) |
| **AWW (25%)** | **8%** | 110.4: (106.5, 114.2) | 110.1: (42.0, 171.0) | 39.9: (39.3, 40.5) | 39.8: (0.0, 136.0) |
| **AWW (50%)** | **8%** | 93.5: (89.7, 97.4) | 93.2: (26.0, 155.0) | 11.9: (11.7, 12.1) | 12.0: (0.0, 55.0) |
| **AWW (100%)** | **8%** | 84.8: (80.9, 88.6) | 84.8: (20.0, 145.0) | 6.1: (5.9, 6.2) | 6.0: (0.0, 31.0) |
| **AWW (10%)** | **16%** | 112.3: (108.4, 116.2) | 112.2: (44.0, 173.0) | 45.4: (44.7, 46.1) | 45.4: (0.0, 146.0) |
| **AWW (25%)** | **16%** | 93.5: (89.7, 97.4) | 93.6: (26.0, 155.0) | 11.9: (11.7, 12.1) | 11.8: (0.0, 54.0) |
| **AWW (50%)** | **16%** | 84.8: (80.9, 88.6) | 84.9: (19.0, 145.0) | 6.1: (5.9, 6.2) | 6.1: (0.0, 32.0) |
| **AWW (100%)** | **16%** | 84.3: (80.4, 88.2) | 84.2: (19.0, 145.0) | 5.9: (5.7, 6.0) | 5.8: (0.0, 31.0) |

***Supplementary Table 2****: Global scenario - $R_0$ = 1.5, Generation Time = 6.0 days.*

| Method (Aircraft Sampling %) | $p_{det}$ | Mean Detection Time (Days) | | EW Local Cases at Time of Detection | |
|---|---|---|---|---|---|
| | | **Means (95% CI)** | **Bootstrap (95% PI)** | **Means (95% CI)** | **Bootstrap (95% PI)** |
| **ICU** | | 132.4: (127.4, 137.4) | 132.5: (42.9, 212.4) | 29.3: (28.8, 29.9) | 29.3: (0.0, 95.0) |
| **AWW (10%)** | **8%** | 137.3: (132.2, 142.4) | 137.3: (51.0, 213.0) | 34.3: (33.4, 35.1) | 34.3: (0.0, 98.0) |
| **AWW (25%)** | **8%** | 133.5: (128.4, 138.5) | 133.4: (48.0, 211.0) | 29.1: (28.5, 29.7) | 29.1: (0.0, 91.0) |
| **AWW (50%)** | **8%** | 117.5: (112.6, 122.5) | 117.5: (32.0, 195.0) | 13.1: (12.9, 13.3) | 13.1: (0.0, 57.0) |
| **AWW (100%)** | **8%** | 106.6: (101.7, 111.5) | 106.6: (22.0, 184.0) | 6.9: (6.8, 7.0) | 6.9: (0.0, 35.0) |
| **AWW (10%)** | **16%** | 134.8: (129.7, 139.8) | 134.8: (48.0, 211.0) | 30.7: (30.1, 31.4) | 30.6: (0.0, 93.0) |
| **AWW (25%)** | **16%** | 117.5: (112.6, 122.5) | 117.5: (31.0, 195.0) | 13.1: (12.9, 13.3) | 13.1: (0.0, 57.0) |
| **AWW (50%)** | **16%** | 106.6: (101.7, 111.5) | 106.7: (22.0, 184.0) | 6.9: (6.8, 7.0) | 6.9: (0.0, 35.0) |
| **AWW (100%)** | **16%** | 106.0: (101.1, 110.9) | 106.1: (22.0, 184.0) | 6.7: (6.6, 6.8) | 6.7: (0.0, 34.0) |

***Supplementary Table 3**: Global scenario - $R_0$ = 1.5, Generation Time = 8.0 days.*

| Method (Aircraft Sampling %) | $p_{det}$ | Mean Detection Time (Days) | | EW Local Cases at Time of Detection | |
|---|---|---|---|---|---|
| | | **Means (95% CI)** | **Bootstrap (95% PI)** | **Means (95% CI)** | **Bootstrap (95% PI)** |
| **ICU** | | 147.8: (142.1, 153.4) | 147.5: (47.0, 233.5) | 26.4: (26.0, 26.9) | 26.5: (0.0, 81.0) |
| **AWW (10%)** | **8%** | 151.2: (145.3, 157.0) | 151.2: (53.0, 234.0) | 28.1: (27.3, 28.8) | 28.0: (0.0, 80.0) |
| **AWW (25%)** | **8%** | 148.4: (142.6, 154.2) | 148.2: (48.0, 231.0) | 25.6: (25.0, 26.1) | 25.4: (0.0, 78.0) |
| **AWW (50%)** | **8%** | 135.3: (129.6, 140.9) | 135.0: (35.0, 220.0) | 15.7: (15.4, 15.9) | 15.6: (0.0, 61.0) |
| **AWW (100%)** | **8%** | 123.2: (117.6, 128.8) | 123.2: (25.0, 208.0) | 9.1: (9.0, 9.3) | 9.1: (0.0, 43.0) |
| **AWW (10%)** | **16%** | 149.6: (143.8, 155.4) | 149.4: (48.0, 231.0) | 26.7: (26.1, 27.3) | 26.6: (0.0, 80.0) |
| **AWW (25%)** | **16%** | 135.3: (129.6, 140.9) | 135.3: (35.0, 220.0) | 15.7: (15.4, 15.9) | 15.6: (0.0, 61.0) |
| **AWW (50%)** | **16%** | 123.2: (117.6, 128.8) | 123.1: (26.0, 208.0) | 9.1: (9.0, 9.3) | 9.1: (0.0, 43.0) |
| **AWW (100%)** | **16%** | 122.3: (116.7, 127.9) | 122.3: (24.0, 207.0) | 8.8: (8.6, 8.9) | 8.7: (0.0, 41.0) |

***Supplementary Table 4**: Global scenario - $R_0$ = 1.5, Generation Time = 10.0 days.*

| Method (Aircraft Sampling %) | $p_{det}$ | Mean Detection Time (Days) | | EW Local Cases at Time of Detection | |
|---|---|---|---|---|---|
| | | **Means (95% CI)** | **Bootstrap (95% PI)** | **Means (95% CI)** | **Bootstrap (95% PI)** |
| **ICU** | | 67.3: (65.5, 69.0) | 67.2: (26.3, 106.5) | 419.0: (411.9, 426.1) | 418.9: (2.0, 1753.0) |
| **AWW (10%)** | **8%** | 57.2: (55.6, 58.9) | 57.2: (29.0, 83.0) | 179.0: (176.2, 181.8) | 179.6: (2.0, 680.0) |
| **AWW (25%)** | **8%** | 52.4: (50.7, 54.0) | 52.3: (24.0, 78.0) | 73.3: (72.2, 74.4) | 74.1: (0.0, 285.0) |
| **AWW (50%)** | **8%** | 45.2: (43.5, 46.9) | 45.3: (17.0, 71.0) | 19.1: (18.8, 19.4) | 19.1: (0.0, 82.0) |
| **AWW (100%)** | **8%** | 41.7: (40.0, 43.4) | 41.8: (13.0, 68.0) | 9.8: (9.7, 10.0) | 9.9: (0.0, 48.0) |
| **AWW (10%)** | **16%** | 53.6: (52.0, 55.3) | 53.6: (25.0, 79.0) | 92.0: (90.4, 93.5) | 92.0: (0.0, 346.0) |
| **AWW (25%)** | **16%** | 45.2: (43.5, 46.9) | 45.2: (17.0, 71.0) | 19.1: (18.8, 19.4) | 19.1: (0.0, 82.0) |
| **AWW (50%)** | **16%** | 41.7: (40.0, 43.4) | 41.8: (13.0, 68.0) | 9.8: (9.7, 10.0) | 9.9: (0.0, 48.0) |
| **AWW (100%)** | **16%** | 41.5: (39.8, 43.2) | 41.4: (13.0, 67.0) | 9.5: (9.3, 9.7) | 9.5: (0.0, 46.0) |

***Supplementary Table 5****: Global scenario - $R_0$ = 2.0, Generation Time = 4.0 days.*

| Method (Aircraft Sampling %) | $p_{det}$ | Mean Detection Time (Days) | | EW Local Cases at Time of Detection | |
|---|---|---|---|---|---|
| | | **Means (95% CI)** | **Bootstrap (95% PI)** | **Means (95% CI)** | **Bootstrap (95% PI)** |
| **ICU** | | 82.4: (80.0, 84.8) | 82.7: (32.9, 129.7) | 280.4: (275.1, 285.7) | 279.2: (1.0, 1127.0) |
| **AWW (10%)** | **8%** | 77.9: (75.6, 80.2) | 77.9: (39.0, 114.0) | 197.7: (194.4, 201.0) | 196.9: (2.0, 733.0) |
| **AWW (25%)** | **8%** | 71.1: (68.8, 73.4) | 71.1: (31.0, 107.0) | 81.2: (79.9, 82.4) | 81.2: (0.0, 314.0) |
| **AWW (50%)** | **8%** | 61.0: (58.6, 63.3) | 61.0: (21.0, 97.0) | 20.9: (20.6, 21.3) | 21.0: (0.0, 90.0) |
| **AWW (100%)** | **8%** | 56.1: (53.7, 58.4) | 56.1: (17.0, 92.0) | 10.8: (10.6, 11.0) | 10.8: (0.0, 52.0) |
| **AWW (10%)** | **16%** | 72.7: (70.4, 75.0) | 72.7: (33.0, 108.0) | 100.3: (98.6, 102.0) | 100.4: (0.0, 385.0) |
| **AWW (25%)** | **16%** | 61.0: (58.6, 63.3) | 61.0: (21.0, 97.0) | 20.9: (20.6, 21.3) | 21.0: (0.0, 90.0) |
| **AWW (50%)** | **16%** | 56.1: (53.7, 58.4) | 56.1: (17.0, 92.0) | 10.8: (10.6, 11.0) | 10.8: (0.0, 51.0) |
| **AWW (100%)** | **16%** | 55.8: (53.5, 58.2) | 55.9: (17.0, 92.0) | 10.4: (10.2, 10.6) | 10.3: (0.0, 49.0) |

***Supplementary Table 6****: Global scenario - $R_0$ = 2.0, Generation Time = 6.0 days.*

| Method (Aircraft Sampling %) | $p_{det}$ | Mean Detection Time (Days) | | EW Local Cases at Time of Detection | |
|---|---|---|---|---|---|
| | | Means (95% CI) | Bootstrap (95% PI) | Means (95% CI) | Bootstrap (95% PI) |
| ICU | | 96.8: (93.8, 99.8) | 96.9: (37.4, 151.9) | 194.1: (190.6, 197.6) | 195.2: (1.0, 699.0) |
| AWW (10%) | 8% | 96.8: (94.0, 99.7) | 96.8: (47.0, 142.0) | 196.2: (193.2, 199.1) | 197.1: (2.0, 698.0) |
| AWW (25%) | 8% | 88.3: (85.4, 91.2) | 88.3: (37.0, 133.0) | 83.6: (82.3, 84.9) | 83.5: (0.0, 322.0) |
| AWW (50%) | 8% | 75.4: (72.4, 78.3) | 75.4: (26.0, 121.0) | 21.1: (20.8, 21.5) | 21.0: (0.0, 91.0) |
| AWW (100%) | 8% | 69.0: (66.1, 72.0) | 69.1: (19.0, 114.0) | 10.9: (10.6, 11.1) | 10.8: (0.0, 51.0) |
| AWW (10%) | 16% | 90.4: (87.5, 93.3) | 90.5: (41.0, 136.0) | 103.4: (101.9, 105.0) | 102.6: (0.0, 391.0) |
| AWW (25%) | 16% | 75.4: (72.4, 78.3) | 75.4: (26.0, 121.0) | 21.1: (20.8, 21.5) | 21.1: (0.0, 91.0) |
| AWW (50%) | 16% | 69.0: (66.1, 72.0) | 69.2: (19.0, 114.0) | 10.9: (10.6, 11.1) | 10.8: (0.0, 51.0) |
| AWW (100%) | 16% | 68.5: (65.6, 71.5) | 68.5: (19.0, 114.0) | 10.1: (9.9, 10.3) | 10.1: (0.0, 48.0) |

***Supplementary Table** 7: Global scenario - $R_0$ = 2.0, Generation Time = 8.0 days.*

| Method (Aircraft Sampling %) | $p_{det}$ | Mean Detection Time (Days) | | EW Local Cases at Time of Detection | |
|---|---|---|---|---|---|
| | | Means (95% CI) | Bootstrap (95% PI) | Means (95% CI) | Bootstrap (95% PI) |
| ICU | | 108.5: (105.0, 112.1) | 108.5: (39.7, 171.6) | 148.9: (146.1, 151.8) | 148.4: (1.0, 497.0) |
| AWW (10%) | 8% | 112.6: (109.1, 116.1) | 112.6: (53.0, 166.0) | 178.8: (175.6, 182.0) | 178.6: (3.0, 501.0) |
| AWW (25%) | 8% | 105.1: (101.6, 108.5) | 105.3: (46.0, 159.0) | 102.1: (100.6, 103.7) | 102.1: (1.0, 368.0) |
| AWW (50%) | 8% | 89.1: (85.6, 92.7) | 89.1: (29.0, 143.0) | 26.5: (26.1, 27.0) | 26.6: (0.0, 112.0) |
| AWW (100%) | 8% | 81.5: (78.0, 85.1) | 81.4: (22.0, 136.0) | 13.6: (13.3, 13.8) | 13.6: (0.0, 61.0) |
| AWW (10%) | 16% | 106.8: (103.4, 110.3) | 106.8: (46.0, 161.0) | 118.9: (117.1, 120.7) | 118.5: (1.0, 405.0) |
| AWW (25%) | 16% | 89.1: (85.6, 92.7) | 89.2: (29.0, 143.0) | 26.5: (26.1, 27.0) | 26.4: (0.0, 112.0) |
| AWW (50%) | 16% | 81.5: (78.0, 85.1) | 81.4: (22.0, 136.0) | 13.6: (13.3, 13.8) | 13.5: (0.0, 62.0) |
| AWW (100%) | 16% | 81.1: (77.6, 84.6) | 81.3: (22.0, 135.0) | 13.0: (12.8, 13.2) | 13.0: (0.0, 59.0) |

***Supplementary Table 8**: Global scenario - $R_0$ = 2.0, Generation Time = 10.0 days.*

| Method (Aircraft Sampling %) | $p_{det}$ | Mean Detection Time (Days) | | EW Local Cases at Time of Detection | |
|---|---|---|---|---|---|
| | | **Means (95% CI)** | **Bootstrap (95% PI)** | **Means (95% CI)** | **Bootstrap (95% PI)** |
| **ICU** | | 66.5: (65.2, 67.8) | 66.5: (25.7, 96.6) | 3039.3: (2992.4, 3086.3) | 3051.5: (5.0, 11109.0) |
| **AWW (10%)** | **8%** | 45.4: (44.2, 46.7) | 45.4: (24.0, 65.0) | 294.5: (289.7, 299.3) | 294.7: (4.0, 1092.0) |
| **AWW (25%)** | **8%** | 41.8: (40.5, 43.1) | 41.8: (20.0, 61.0) | 121.9: (119.9, 123.9) | 122.1: (1.0, 468.0) |
| **AWW (50%)** | **8%** | 36.6: (35.3, 37.9) | 36.6: (15.0, 56.0) | 32.7: (32.2, 33.3) | 32.9: (0.0, 133.0) |
| **AWW (100%)** | **8%** | 33.9: (32.6, 35.2) | 33.9: (12.0, 53.0) | 16.5: (16.3, 16.8) | 16.7: (0.0, 77.0) |
| **AWW (10%)** | **16%** | 42.7: (41.4, 44.0) | 42.7: (21.0, 62.0) | 152.4: (149.9, 154.9) | 153.6: (1.0, 580.0) |
| **AWW (25%)** | **16%** | 36.6: (35.3, 37.9) | 36.6: (15.0, 56.0) | 32.7: (32.2, 33.3) | 32.6: (0.0, 134.0) |
| **AWW (50%)** | **16%** | 33.9: (32.6, 35.2) | 33.9: (12.0, 53.0) | 16.5: (16.3, 16.8) | 16.5: (0.0, 76.0) |
| **AWW (100%)** | **16%** | 33.8: (32.5, 35.1) | 33.8: (12.0, 53.0) | 16.1: (15.8, 16.4) | 16.2: (0.0, 74.0) |

***Supplementary Table 9****: Global scenario - $R_0$ = 2.5, Generation Time = 4.0 days.*

| Method (Aircraft Sampling %) | $p_{det}$ | Mean Detection Time (Days) | | EW Local Cases at Time of Detection | |
|---|---|---|---|---|---|
| | | **Means (95% CI)** | **Bootstrap (95% PI)** | **Means (95% CI)** | **Bootstrap (95% PI)** |
| **ICU** | | 77.6: (75.9, 79.4) | 77.7: (31.7, 115.6) | 1336.6: (1314.0, 1359.2) | 1340.1: (3.0, 4475.1) |
| **AWW (10%)** | **8%** | 60.7: (59.0, 62.4) | 60.8: (31.0, 88.0) | 300.6: (295.6, 305.6) | 300.0: (4.0, 1138.0) |
| **AWW (25%)** | **8%** | 55.7: (54.0, 57.4) | 55.6: (26.0, 82.0) | 121.9: (119.9, 123.9) | 121.8: (1.0, 464.0) |
| **AWW (50%)** | **8%** | 48.3: (46.6, 50.1) | 48.3: (19.0, 75.0) | 31.7: (31.1, 32.2) | 31.7: (0.0, 131.0) |
| **AWW (100%)** | **8%** | 44.7: (42.9, 46.4) | 44.6: (15.0, 72.0) | 16.0: (15.8, 16.3) | 16.1: (0.0, 74.0) |
| **AWW (10%)** | **16%** | 57.0: (55.3, 58.7) | 57.0: (28.0, 84.0) | 153.5: (151.1, 155.8) | 152.9: (1.0, 576.0) |
| **AWW (25%)** | **16%** | 48.3: (46.6, 50.1) | 48.3: (19.0, 75.0) | 31.7: (31.1, 32.2) | 31.7: (0.0, 131.0) |
| **AWW (50%)** | **16%** | 44.7: (42.9, 46.4) | 44.7: (15.0, 72.0) | 16.0: (15.8, 16.3) | 16.1: (0.0, 73.0) |
| **AWW (100%)** | **16%** | 44.5: (42.7, 46.2) | 44.5: (15.0, 71.0) | 15.5: (15.3, 15.8) | 15.5: (0.0, 71.0) |

***Supplementary Table 10****: Global scenario - $R_0$ = 2.5, Generation Time = 6.0 days.*

| Method (Aircraft Sampling %) | $p_{det}$ | Mean Detection Time (Days) | | EW Local Cases at Time of Detection | |
|---|---|---|---|---|---|
| | | Means (95% CI) | Bootstrap (95% PI) | Means (95% CI) | Bootstrap (95% PI) |
| **ICU** | | 87.6: (85.3, 89.8) | 87.5: (33.7, 131.5) | 766.5: (753.4, 779.5) | 768.2: (2.0, 2413.0) |
| **AWW (10%)** | **8%** | 74.6: (72.5, 76.7) | 74.7: (38.0, 108.0) | 289.9: (285.0, 294.7) | 289.1: (4.0, 1083.0) |
| **AWW (25%)** | **8%** | 68.4: (66.2, 70.5) | 68.4: (32.0, 102.0) | 118.8: (116.9, 120.7) | 118.5: (1.0, 448.0) |
| **AWW (50%)** | **8%** | 58.9: (56.7, 61.1) | 58.9: (22.0, 92.0) | 30.2: (29.7, 30.7) | 30.1: (0.0, 126.0) |
| **AWW (100%)** | **8%** | 54.4: (52.3, 56.6) | 54.5: (18.0, 88.0) | 15.4: (15.2, 15.7) | 15.4: (0.0, 68.0) |
| **AWW (10%)** | **16%** | 69.8: (67.7, 72.0) | 69.9: (33.0, 103.0) | 147.8: (145.4, 150.1) | 148.0: (1.0, 576.0) |
| **AWW (25%)** | **16%** | 58.9: (56.7, 61.1) | 58.9: (22.0, 92.0) | 30.2: (29.7, 30.7) | 30.3: (0.0, 125.0) |
| **AWW (50%)** | **16%** | 54.4: (52.3, 56.6) | 54.4: (18.0, 88.0) | 15.4: (15.2, 15.7) | 15.4: (0.0, 69.0) |
| **AWW (100%)** | **16%** | 54.1: (51.9, 56.3) | 54.0: (17.0, 87.0) | 14.7: (14.5, 15.0) | 14.8: (0.0, 66.0) |

***Supplementary Table 11***: *Global scenario - $R_0$ = 2.5, Generation Time = 8.0 days.*

| Method (Aircraft Sampling %) | $p_{det}$ | Mean Detection Time (Days) | | EW Local Cases at Time of Detection | |
|---|---|---|---|---|---|
| | | Means (95% CI) | Bootstrap (95% PI) | Means (95% CI) | Bootstrap (95% PI) |
| **ICU** | | 95.7: (93.1, 98.3) | 95.7: (38.1, 145.8) | 565.9: (557.1, 574.7) | 564.9: (2.0, 1792.0) |
| **AWW (10%)** | **8%** | 87.8: (85.3, 90.3) | 87.7: (44.0, 127.0) | 344.2: (338.5, 349.9) | 342.6: (5.0, 1295.0) |
| **AWW (25%)** | **8%** | 80.3: (77.7, 82.8) | 80.1: (36.0, 119.0) | 138.7: (136.6, 140.8) | 138.1: (1.0, 523.0) |
| **AWW (50%)** | **8%** | 69.0: (66.4, 71.6) | 68.9: (25.0, 109.0) | 35.4: (34.8, 36.0) | 35.4: (0.0, 146.0) |
| **AWW (100%)** | **8%** | 63.5: (60.9, 66.1) | 63.5: (19.0, 103.0) | 17.9: (17.6, 18.3) | 18.0: (0.0, 80.0) |
| **AWW (10%)** | **16%** | 82.2: (79.6, 84.7) | 82.2: (39.0, 122.0) | 174.4: (171.7, 177.1) | 173.7: (2.0, 668.0) |
| **AWW (25%)** | **16%** | 69.0: (66.4, 71.6) | 69.1: (25.0, 109.0) | 35.4: (34.8, 36.0) | 35.3: (0.0, 146.0) |
| **AWW (50%)** | **16%** | 63.5: (60.9, 66.1) | 63.6: (19.0, 103.0) | 17.9: (17.6, 18.3) | 18.0: (0.0, 79.0) |
| **AWW (100%)** | **16%** | 63.2: (60.6, 65.8) | 63.1: (19.0, 103.0) | 17.4: (17.1, 17.7) | 17.4: (0.0, 78.0) |

***Supplementary Table 12***: *Global scenario - $R_0$ = 2.5, Generation Time = 10.0 days.*

| Method (Aircraft Sampling %) | $p_{det}$ | Mean Detection Time (Days) | | EW Local Cases at Time of Detection | |
|---|---|---|---|---|---|
| | | **Means (95% CI)** | **Bootstrap (95% PI)** | **Means (95% CI)** | **Bootstrap (95% PI)** |
| **ICU** | | 60.9: (59.8, 62.0) | 60.9: (26.1, 88.0) | 12616.2: (12396.8, 12835.6) | 12628.4: (12.0, 54310.0) |
| **AWW (10%)** | **8%** | 39.1: (38.0, 40.1) | 39.1: (21.0, 55.0) | 452.8: (446.2, 459.5) | 452.8: (8.0, 1669.0) |
| **AWW (25%)** | **8%** | 36.1: (35.0, 37.2) | 36.1: (18.0, 52.0) | 189.4: (186.5, 192.2) | 188.8: (2.0, 716.0) |
| **AWW (50%)** | **8%** | 31.7: (30.7, 32.8) | 31.7: (14.0, 48.0) | 50.7: (49.9, 51.5) | 50.7: (0.0, 206.0) |
| **AWW (100%)** | **8%** | 29.6: (28.5, 30.7) | 29.6: (12.0, 46.0) | 26.0: (25.6, 26.4) | 26.2: (0.0, 115.0) |
| **AWW (10%)** | **16%** | 36.9: (35.8, 37.9) | 36.9: (19.0, 53.0) | 235.6: (232.0, 239.2) | 236.4: (3.0, 869.0) |
| **AWW (25%)** | **16%** | 31.7: (30.7, 32.8) | 31.7: (14.0, 48.0) | 50.7: (49.9, 51.5) | 50.8: (0.0, 207.0) |
| **AWW (50%)** | **16%** | 29.6: (28.5, 30.7) | 29.6: (12.0, 46.0) | 26.0: (25.6, 26.4) | 26.2: (0.0, 116.0) |
| **AWW (100%)** | **16%** | 29.4: (28.4, 30.5) | 29.4: (12.0, 46.0) | 24.7: (24.3, 25.1) | 24.7: (0.0, 110.0) |

***Supplementary Table 13****: Global scenario - $R_0$ = 3.0, Generation Time = 4.0 days.*

| Method (Aircraft Sampling %) | $p_{det}$ | Mean Detection Time (Days) | | EW Local Cases at Time of Detection | |
|---|---|---|---|---|---|
| | | **Means (95% CI)** | **Bootstrap (95% PI)** | **Means (95% CI)** | **Bootstrap (95% PI)** |
| **ICU** | | 74.9: (73.4, 76.3) | 74.8: (31.4, 105.7) | 4648.2: (4578.9, 4717.6) | 4651.6: (7.0, 14535.0) |
| **AWW (10%)** | **8%** | 51.6: (50.2, 53.0) | 51.6: (27.0, 74.0) | 432.5: (425.4, 439.6) | 431.7: (6.0, 1624.0) |
| **AWW (25%)** | **8%** | 47.5: (46.0, 48.9) | 47.4: (23.0, 69.0) | 175.4: (172.6, 178.2) | 175.2: (1.0, 667.0) |
| **AWW (50%)** | **8%** | 41.4: (40.0, 42.9) | 41.5: (17.0, 63.0) | 45.8: (45.0, 46.5) | 45.8: (0.0, 190.0) |
| **AWW (100%)** | **8%** | 38.4: (37.0, 39.9) | 38.4: (14.0, 60.0) | 23.2: (22.8, 23.6) | 23.2: (0.0, 104.0) |
| **AWW (10%)** | **16%** | 48.5: (47.1, 49.9) | 48.6: (24.0, 70.0) | 219.6: (215.9, 223.3) | 216.8: (2.0, 819.0) |
| **AWW (25%)** | **16%** | 41.4: (40.0, 42.9) | 41.5: (17.0, 63.0) | 45.8: (45.0, 46.5) | 46.0: (0.0, 192.0) |
| **AWW (50%)** | **16%** | 38.4: (37.0, 39.9) | 38.4: (14.0, 60.0) | 23.2: (22.8, 23.6) | 23.2: (0.0, 104.0) |
| **AWW (100%)** | **16%** | 38.2: (36.8, 39.7) | 38.2: (14.0, 60.0) | 22.2: (21.8, 22.6) | 22.3: (0.0, 101.0) |

***Supplementary Table 14****: Global scenario - $R_0$ = 3.0, Generation Time = 6.0 days.*

| Method (Aircraft Sampling %) | $p_{det}$ | Mean Detection Time (Days) | | EW Local Cases at Time of Detection | |
|---|---|---|---|---|---|
| | | Means (95% CI) | Bootstrap (95% PI) | Means (95% CI) | Bootstrap (95% PI) |
| **ICU** | | 83.9: (82.1, 85.8) | 83.9: (35.3, 119.8) | 2259.9: (2227.3, 2292.5) | 2259.8: (4.0, 6465.0) |
| **AWW (10%)** | **8%** | 62.7: (61.0, 64.4) | 62.7: (33.0, 90.0) | 398.7: (392.4, 404.9) | 399.2: (6.0, 1484.0) |
| **AWW (25%)** | **8%** | 57.7: (55.9, 59.4) | 57.6: (28.0, 85.0) | 163.9: (161.2, 166.6) | 164.7: (1.0, 616.0) |
| **AWW (50%)** | **8%** | 50.0: (48.2, 51.8) | 50.0: (20.0, 77.0) | 41.9: (41.3, 42.6) | 41.8: (0.0, 178.0) |
| **AWW (100%)** | **8%** | 46.3: (44.5, 48.0) | 46.2: (16.0, 73.0) | 21.0: (20.6, 21.4) | 21.2: (0.0, 97.0) |
| **AWW (10%)** | **16%** | 58.8: (57.1, 60.6) | 58.8: (29.0, 86.0) | 203.2: (199.9, 206.6) | 204.2: (2.0, 776.0) |
| **AWW (25%)** | **16%** | 50.0: (48.2, 51.8) | 50.0: (20.0, 77.0) | 41.9: (41.3, 42.6) | 42.1: (0.0, 178.0) |
| **AWW (50%)** | **16%** | 46.3: (44.5, 48.0) | 46.2: (16.0, 73.0) | 21.0: (20.6, 21.4) | 20.9: (0.0, 95.0) |
| **AWW (100%)** | **16%** | 46.1: (44.3, 47.9) | 46.1: (16.0, 73.0) | 20.3: (19.9, 20.7) | 20.3: (0.0, 94.0) |

***Supplementary Table 15****: Global scenario - $R_0$ = 3.0, Generation Time = 8.0 days.*

| Method (Aircraft Sampling %) | $p_{det}$ | Mean Detection Time (Days) | | EW Local Cases at Time of Detection | |
|---|---|---|---|---|---|
| | | Means (95% CI) | Bootstrap (95% PI) | Means (95% CI) | Bootstrap (95% PI) |
| **ICU** | | 90.4: (88.2, 92.5) | 90.2: (37.5, 132.7) | 1508.8: (1487.6, 1530.1) | 1500.2: (4.0, 4396.0) |
| **AWW (10%)** | **8%** | 73.2: (71.2, 75.3) | 73.3: (38.0, 105.0) | 454.8: (447.1, 462.5) | 456.2: (7.0, 1709.0) |
| **AWW (25%)** | **8%** | 67.1: (65.1, 69.2) | 67.2: (32.0, 99.0) | 182.4: (179.6, 185.3) | 181.7: (2.0, 684.0) |
| **AWW (50%)** | **8%** | 58.1: (56.0, 60.2) | 58.1: (22.0, 90.0) | 47.1: (46.4, 47.9) | 47.1: (0.0, 194.0) |
| **AWW (100%)** | **8%** | 53.6: (51.5, 55.7) | 53.7: (18.0, 86.0) | 23.7: (23.3, 24.1) | 23.7: (0.0, 104.0) |
| **AWW (10%)** | **16%** | 68.6: (66.6, 70.7) | 68.6: (33.0, 101.0) | 229.5: (225.7, 233.4) | 229.4: (2.0, 849.0) |
| **AWW (25%)** | **16%** | 58.1: (56.0, 60.2) | 58.0: (22.0, 90.0) | 47.1: (46.4, 47.9) | 47.1: (0.0, 195.0) |
| **AWW (50%)** | **16%** | 53.6: (51.5, 55.7) | 53.6: (19.0, 86.0) | 23.7: (23.3, 24.1) | 23.6: (0.0, 104.0) |
| **AWW (100%)** | **16%** | 53.4: (51.3, 55.5) | 53.4: (18.0, 86.0) | 22.8: (22.4, 23.2) | 22.7: (0.0, 99.0) |

***Supplementary Table 16****: Global scenario - $R_0$ = 3.0, Generation Time = 10.0 days.*
